# Clinical Validation of Digital PCR-based ctDNA detection for risk stratification in residual triple negative breast cancer: TRICIA trial results

**DOI:** 10.1101/2025.07.02.25330747

**Authors:** Talia Roseshter, Anna Klemantovich, Josiane Lafleur, Cathy Lan, Luca Cavallone, Kathryn Bozek, Juliet Guay, Oluwadara Elebute, Sarah Jenna, Rodney Ouellette, Suzan McNamara, Jean-Francois Boileau, Manuela Pelmus, Muriel Brackstone, Rossanna Pezo, Terry Ng, Adriana Aguilar-Mahecha, Mark Basik

**Author notes:** Corresponding Author: **Mark Basik,****, Dept of Oncology, Jewish General Hospital, 3755 Cote Ste Catherine, Montreal, Qc, H3T1E2, Canada**. A. Aguilar-Mahecha and M.Basik contributed equally as co-last authors. **Conflict of Interest Statement:** The authors declare no potential conflicts of interest.

## Abstract

Triple-negative breast cancer (TNBC) patients who have residual tumor at surgery (non-pathological complete response or non-pCR) after neoadjuvant chemotherapy (NAC) have a poor prognosis. In these cases, adjuvant chemotherapy with capecitabine improves disease-free survival in ∼15% of patients. Identifying those who would benefit or not from such additional therapy remains a critical need. Circulating tumor DNA (ctDNA), a plasma-based biomarker, provides real-time insights into disease and treatment progression. We previously demonstrated that ctDNA detection after NAC and before surgery signals poor prognosis. In the TRICIA trial, 92 patients with non-pCR provided plasma before surgery and after NAC (T1), after surgery (T2), during adjuvant capecitabine therapy (T3) and late after surgery following completion of adjuvant treatment (T4). The sensitivity, specificity, and predictive values of a tumor-informed digital-droplet-based ctDNA detection assay were measured with a median follow-up of 38 months. ctDNA was detected in 97% of patients before clinical relapse. We confirmed that the lack of detection of ctDNA at the post-NAC pre-operative (T1) time point is highly prognostic, with 95% distant-disease relapse free survival. The other time points were not as strongly prognostic. The detection of ctDNA in patients with significant residual tumor (Residual Cancer Burden 2 or 3) was also highly prognostic and our test performed with 100% sensitivity and 100% specificity in RCB 3 patients. Although the extent of residual disease was correlated with the Fractional Abundance of individual variants, the effect of surgery on ctDNA detectability was not significant except for RCB 3 cases, in which large amounts of residual disease were removed. We measured 3 time points before, during and after capecitabine treatment and found that capecitabine treatment was associated with clearance of ctDNA in 41% of cases, and clearance (from detection to non-detection) was associated with good prognosis. These findings suggest that ctDNA testing using ddPCR assays in an academic hospital-based context can reliably identify a very low-risk group of non-pCR TNBC patients, and this personalized approach is ready for prospective testing for clinical utility in TNBC patients who have undergone NAC and require additional chemotherapy.

## Introduction

Triple-negative breast cancer (TNBC) is the most aggressive form of breast cancer and accounts for about 15-20% of all breast cancers^1^. The term “triple-negative” refers to the lack of protein expression of the three most common biomarkers in the treatment of breast cancer: estrogen receptors (ER), progesterone receptors (PR), and human epidermal growth factor receptor 2 (HER2). The lack of these receptors makes TNBC unresponsive to common hormonal therapies or HER2-targeted treatments leaving chemotherapy as the mainstay of treatment. In early-stage TNBC, neoadjuvant chemotherapy (NAC) is used to reduce the size of the tumor prior to surgery. The extent of response to NAC is a strong prognostic marker; patients with residual tumor at the time of surgery (non-pathological complete response or non-pCR) have a significantly higher risk of relapse and death. About half of TNBC patients treated with neoadjuvant chemotherapy (NAC) have residual disease and 35% of these patients show disease recurrence within 2 years^2^. The residual cancer burden (RCB) score, which measures the extent of residual disease in the tumor and lymph nodes, serves as an additional prognostic tool to stratify recurrence risk^3^. The CREATE-X clinical trial^4^ demonstrated that the addition of adjuvant chemotherapy (capecitabine) results in improved survival in patients with non-pCR. However, only about 15% of patients derive benefit from this additional chemotherapy^4^, suggesting that the majority, around 85% of these patients, may be exposed to unnecessary toxicity with no improvement in outcomes. Identifying those patients that can truly benefit from further adjuvant chemotherapy is an unmet need in the care of patients with TNBC.

Recently, plasma-based biomarkers or liquid biopsies have emerged as possible cancer biomarkers^5^. One such biomarker is circulating tumor DNA (ctDNA). ctDNA are short tumor-derived fragments of DNA that circulate freely in serum and plasma^6^ and represent a small proportion of total cell free DNA (cfDNA)^7^. Measuring ctDNA from serial blood samples can provide real time information about the disease and treatment progression^8^. The measurement of ctDNA is ultra-specific (detecting DNA variants present only in tumor cells) and very sensitive (with levels of detection of ctDNA in the range of 0.1% - 10% of total circulating free DNA^9^). Previous work has shown that the detection of ctDNA after surgical resection of the primary breast tumor indicates poor prognosis, suggesting the presence of minimal residual disease (MRD) in the body^10^. The detection of the minute quantities of ctDNA indicative of MRD require tumor-bespoke or tumor-informed assays, in which personalized assays are developed based on DNA sequencing of each tumor. Using such a tumor bespoke assay developed in an academic setting, we recently showed that the presence of detectable ctDNA after chemotherapy but prior to surgery was a reliable indicator of poor prognosis^11^.

The TRICIA trial (NCT04874064) focussed on TNBC patients with non-pCR, with the aim to investigate tissue and plasma-based biomarkers associated with resistance to chemotherapy in this disease. A key part of its objectives is to both validate and extend our previous findings of the strong prognostic value of pre-operative ctDNA testing. In addition, we included bloods collected at 3 different post-operative time points. During the study, the standard of care changed, and patients received adjuvant capecitabine. The analysis of serial plasma samples in these patients receiving capecitabine forms the largest cohort of ctDNA taken prior to, during and after adjuvant capecitabine treatment in TNBC patients. In the current study, we. validated the very strong prognostic value of the post-NAC, pre-operative time point (T1), as well as demonstrated the prognostic value of ctDNA clearance during capecitabine treatment.

## Methods

### Patient recruitment

Female patients (individuals assigned female at birth) were invited to participate in the TRIple Negative Breast Cancer markers in Liquid Biopsies using Artificial Intelligence trial (TRICIA trial, NCT04874064) if they had triple negative breast cancer and had residual tumor following standard of care neoadjuvant chemotherapy. Sixty-nine patients were recruited in 3 different centers in Canada including the Jewish General Hospital, Ottawa Hospital Research Institute, and London Health Sciences Center between 2020-2022. The last participant was enrolled in August 2022. This study was approved by the REB from the Jewish General Hospital (Protocol MP-05-2020-1813), and the Ontario Cancer Research Ethics Board (OCREB, CTO Project ID: 1934).

To expand our TRICIA cohort, we also included 48 patients with the same inclusion criteria from the JGH breast biobank (JGH REB protocol 05-006). In accordance with the Declaration of Helsinki; all patients provided written informed consent. A total of 117 patients were consented and 92 were included in this study (Consort Diagram **Supplementary Figure 1**). For one of the patients, there were two different TNBC tumors (one from each breast) that were sequenced and ctDNA assays developed, the results were included as independent ctDNA patient results for all statistical analyses. Patient clinical outcomes (relapse-free and overall survival) were concealed until the time of analysis.

### Blood collection and processing

Serial blood samples were collected at different time points: post-NAC but prior to surgery (T1), early post-op (T2), and late post-op or post-adjuvant treatment (T4). For 67 patients receiving capecitabine, T2 bloods were taken prior to capecitabine treatment, a T3 collection was added during capecitabine and T4 bloods for these patients were always collected after completion of adjuvant capecitabine treatment. For each time point, blood was collected in 6 mL k-EDTA collection tubes and processed within 2 hours of collection following standard operating procedures. The samples were centrifuged at 2500 rpm for 15 minutes at room temperature. Plasma and buffy coats were aliquoted into barcoded 1.5 mL tubes and stored at-80°C. Prior to cell free DNA extraction, aliquots were thawed and subjected to a second centrifugation at 10,000 rpm at room temperature.

Commercial pooled human plasma (blood derived, IPLAWBK2E1000ML) was purchased from, Innovative Research (Lot#36336), aliquoted and frozen at-80°C. Cell free DNA was extracted and used as normal plasma control to measure the background fluorescence and to establish variant allele frequency (VAF) detection thresholds for all tested assays.

Cell-free DNA (cfDNA) extraction was performed from 2 mL of plasma using our modified hybrid JGH extraction protocol as previously reported^11, 12^. DNA was extracted from buffy coats using the QIAamp DNA Mini Kit (Qiagen; Cat# 51304).

### Tumor whole exome sequencing and variant calling

Formalin-fixed paraffin-embedded (FFPE) samples of residual tumors or diagnostic biopsies were used for whole exome sequencing. 10-micron sections were used for DNA extraction using the QIAamp DNA FFPE Advanced UNG Kit (Qiagen, Cat #: 56704). Macrodissection was performed to enrich for tumor cells when required. All extractions were performed on tissue containing at least 50% tumor cellularity confirmed by a pathologist (MP).

200 ng of gDNA from tumor and lymphocytes were used for library preparation at the Institute for Research in Immunology and Cancer’s Genomics Platform (IRIC). Libraries were prepared using the SureSelect XT with enzymatic fragmentation library prep kit (Agilent) according to the manufacturer’s instructions. Exome capture was performed using SureSelect Human All Exon v7 library prep kit (Agilent). Libraries were quantified by QuBit and BioAnalyzer DNA1000. All libraries were diluted to 10 nM and normalized by qPCR using the KAPA library quantification kit (KAPA; Cat no. KK4973). Libraries were pooled to equimolar concentration. Sequencing was performed at the McGill Genome Center on the Illumina NovaSeq S4 200 cycles flowcell (2×100bp).

Preprocessing, including removal of the molecular barcode, adapter sequences and low-quality sequences was performed using fastp. Reads were then aligned to the hg37 genome using the Burrows-Wheeler Aligner bwa-mem tool. The variants were called using the GATK4 best practices workflow for somatic short variant discovery (https://gatk.broadinstitute.org/hc/en-us/articles/360035535912-Data-pre-processing-for-variant-discovery). Additionally, a panel of normal sequences using all lymphocyte germline data from the study was created to capture technical artifacts. The GATK4 best practices workflow was followed up to the FilterMutectCalls step (https://gatk.broadinstitute.org/hc/en-us/articles/360035894731-Somatic-short-variant-discovery-SNVs-Indels). Finally, variants were annotated using snpeff tool (http://pcingola.github.io/SnpEff/). Mutect2 vcf output includes a two log-odd ratios confidence score indicating the likelihood that the tumor is present. BCFtools was used to filter for only variants that passed the GATK4 tool scoring (TLOD > 6.3). The data were then passed through the following filters: a mapping quality above 40, a read depth of variant allele frequency above 10 reads, and the variant being absent from the panel of normal sequences. The top 5 somatic variants per patient were selected for ddPCR assay development based on the highest TLOD score and the highest VAF. Only variants with VAFs greater than the median VAF values of all filtered variants were considered for each patient.

### Development of digital droplet PCR assays and measurements

Five ddPCR assays were developed per patient using a set of compatible primers and probes for each variant designed and ordered from Integrated DNA Technologies (IDT) (see **Supplementary Table 1**). ddPCR conditions were optimized to determine the optimal annealing temperature for each assay, ranging from 52°C to 62°C, using the C1000 Touch Thermal Cycler with a 96-deep well reaction module (Cat# 80224, Bio-Rad). Each ddPCR assay was validated using DNA extracted from the respective tumor.

20 ng of cfDNA was used for targeted pre-amplification reactions with the SsoAdvanced PreAmp Supermix (Cat#1725160, BioRad) according to the manufacturer’s instructions. Detection of mutant variants was performed using the Droplet Digital PCR system (QX200 Droplet Generator, Cat# 1864002; PX1 PCR Plate Sealer, Cat# 1814000; C1000 Touch Thermal Cycler with 96-deep well reaction module, Cat# 80224; QX200 Droplet Reader, Cat# 17005228; Bio-Rad) and ddPCR Supermix for Probes (no UTP) (Cat# Q33216; Bio-Rad). The final concentration of the primer/probe mix was 900 nM/250 nM. Preamplified DNA was diluted 100-fold to achieve optimal resolution. For ddPCR reactions, 8 µl of diluted preamplified DNA per well was used. The thermal cycling protocol was: 10 min at 95 °C; (30 sec at 94 °C, 1 min at Ta °C) × 50 cycles; 10 min at 95 °C; hold at 4 °C. QX Manager 2.2 Standard Edition was used for cluster determination and VAF calculation.

All samples were run in triplicate and each run included normal plasma control cfDNA, the patient’s lymphocyte DNA to control for clonal hematopoiesis of indeterminate potential (CHIP) as well as tumor DNA as a positive control. For some patients, additional positive controls included pre-NAC treatment cfDNA samples and cfDNA samples from the metastatic stage. All controls were run in triplicate.

A variant was considered detectable if the variant allele frequency (VAF = proportion of mutant alleles/non-mutant alleles) was greater than at least 2X the standard deviation (SD) of the VAF of the normal plasma controls^11^ and there were at least three positive droplets across replicates.

Corrected fractional abundance was calculated by subtracting the threshold of detection (variant-specific threshold of detection) from the average fractional abundance of a given time point sample of each variant (in triplicate).

## Statistical Analysis

Kaplan-Meier (KM) survival curve analysis: relapse free survival (RFS) was calculated from time of surgery to time of relapse or last follow up. Overall survival (OS) was calculated from time of diagnosis to time of death or last follow up. KM curves were generated for RFS and OS using Graph Pad Prism (RRID:SCR_002798).

KM curves were analyzed for statistical differences between ctDNA status, capecitabine treatment, and RCB score analyses using log rank (Mantel-Cox) tests for significance and log rank tests for calculating Hazard Ratio with confidence intervals in Graph Pad Prism (RRID:SCR_002798).

Bar graphs and violin plots were analyzed by unpaired two-tailed t-tests for comparing cFA between two groups (relapse vs non-relapse, ctDNA positive vs negative, RCB scores, and time points) or one-way ANOVA for comparing cFA between three or more groups (RCB scores and time points). Paired two-tailed t-tests were used for comparing changes in cFA values of matched samples. To evaluate the correlation between Tumor cFA and Tumor VAF, the parametric Pearson correlation (r) test was used. Fisher’s exact test was used for comparing proportions (stacked bar graphs associated with KM plots). All statistical analyses were performed in GraphPad Prism (RRID:SCR_002798).

Performance metrics were calculated as follows:

Positive Predictive Value = ctDNA+ relapses/(ctDNA+ relapses **+** ctDNA+ non-relapses)

Negative Predictive Value = ctDNA-non-relapses/(ctDNA-non-relapses **+** ctDNA-relapses)

Sensitivity = ctDNA+ relapses/(ctDNA+ relapses **+** ctDNA-relapses)

Specificity = ctDNA-relapses/(ctDNA-relapses **+** ctDNA+ non-relapses)

Local relapse is defined as a recurrence in the breast tissue or axillary lymph nodes on the same side as the primary tumor. Distant relapse is a recurrence at a distant site away from the original breast tumor.

### RCB score Calculations

RCB scores were evaluated using the criteria described by the MD Anderson Cancer Center (https://www.mdanderson.org/content/dam/mdanderson/documents/for-physicians/clinical-calculators/PLM_protocol_RCB_calculators_BRO.pdf.)

### Whole Exome Sequencing Data

Data will be made available upon request.

## Results

### Patient Characteristics

A total of 117 TNBC patients with non-pathological complete response following neoadjuvant chemotherapy were part of the TRICIA study cohort. 92 patients with high quality tumor WES data and blood collected at least 1 of the 4 pre-specified time points were included in this ctDNA study (**Supplementary Figure 1**). Blood was collected at the following time points: T1 post-NAC but prior to surgery (median = 0.43 months before surgery, range 0 – 4.6 months), T2 early post-surgery (median = 1.1 months, range 0.03 – 4.1 months), T3 during adjuvant treatment (median = 3.8 months, range 1.4 – 8.0 months), and T4 late post-op or at the end of adjuvant treatment (median = 8.7 months, range 3.5 – 15.0 months) (**Figure 1A**). The T3 time point was only collected in patients who received adjuvant capecitabine.

**Figure 1.**
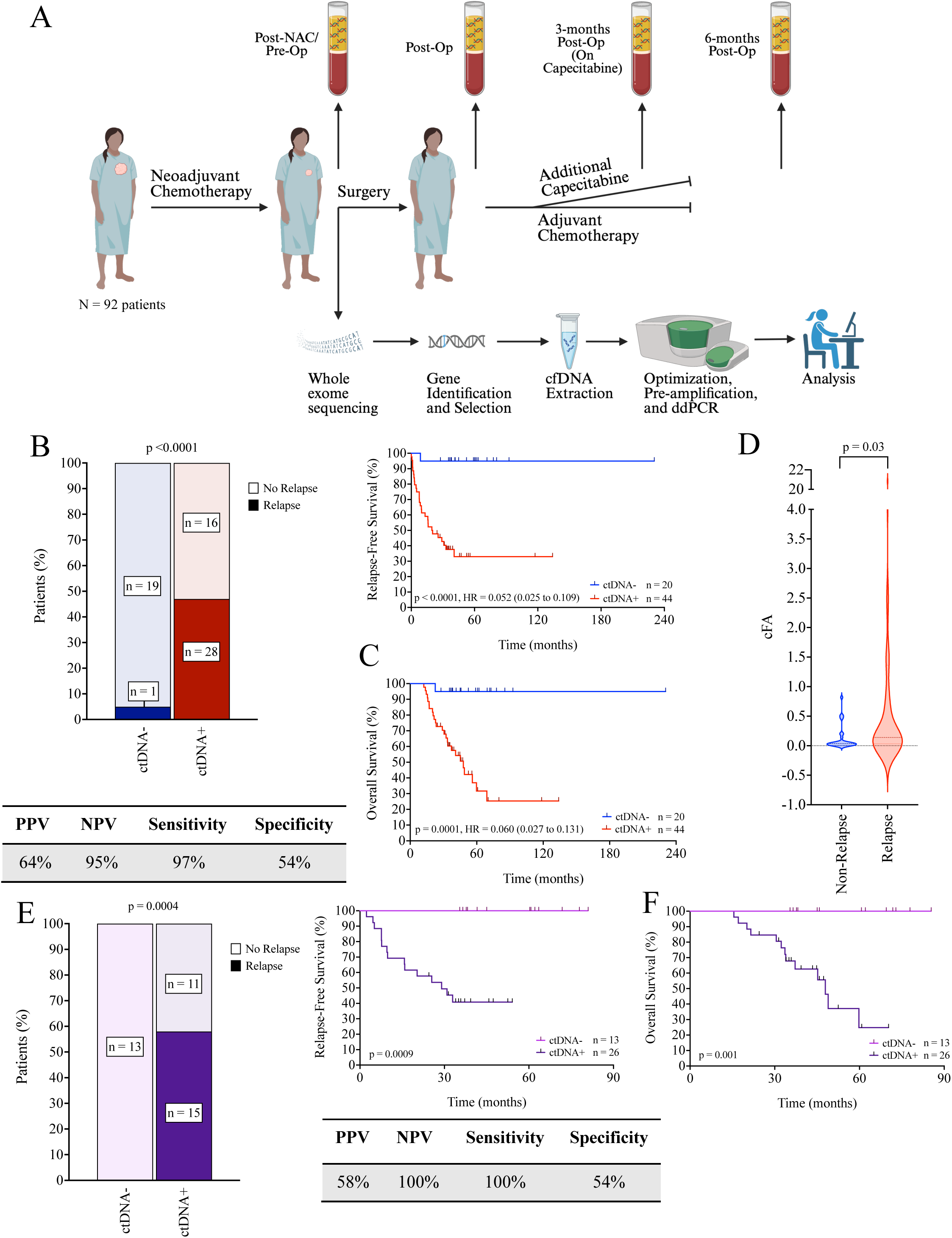
TRICIA overview and prognostic performance of ctDNA detection at the post-neoadjuvant, pre-surgical time point (T1). **(A)** Overview of the TRICIA project sample collection and analysis as described in the METHODS section. **(B)** Proportions of patients with detectable (ctDNA+) and undetectable (ctDNA-) ctDNA at T1 who experienced relapse or remained relapse-free. Statistical comparison of proportions was performed using Fisher’s exact test. Diagnostic performance table includes the PPV, NPV, sensitivity, and specificity of ctDNA status at T1 for predicting relapse. RFS was assessed by Kaplan–Meier (KM) analysis, stratified by ctDNA status. P-value was calculated using the log-rank test and associated hazard ratios. **(C)** KM analysis of OS in the same cohort, stratified by T1 ctDNA status. P-value was calculated using the log-rank test and associated hazard ratios. **(D)** Violin plot comparing the mean fractional abundance of mutant alleles at T1 between relapsed and non-relapsed patients. P-value was calculated using an unpaired two-tailed Student’s t-test. **(E)** Subgroup analysis of patients who received additional adjuvant capecitabine. Proportions of patients with detectable (ctDNA+) and undetectable (ctDNA-) ctDNA at T1 who experienced relapse or remained relapse-free. A diagnostic performance table includes the PPV, NPV, sensitivity, and specificity of ctDNA status at T1 for predicting relapse. RFS was assessed by KM analysis, stratified by ctDNA status. P-value was calculated using the log-rank test and associated hazard ratios. **(F)** KM analysis of OS in the capecitabine-treated subgroup stratified by T1 ctDNA status. P-value was calculated using the log-rank test and associated hazard ratios.

The clinical characteristics of this cohort are found in **Table 1**. The median age range of the patients was 55.5 years old (range: 28 – 79), 92% had stage II or III disease at the time of diagnosis. Most patients received standard Adriamycin/Cyclophosphamide/Paclitaxel neoadjuvant chemotherapy (87%) with carboplatin added in 41% of cases and Atezolizumab in 10% of cases. The median follow-up of these patients was 38.2 months (12.2 – 236.4 months), At the time of data cut-off, 37 (40%) patients had relapsed (31 distant and 6 local relapses) and 27 patients had died. 61 of 92 patients (66%) received adjuvant capecitabine and among these, 20 patients (33%) experienced relapse. Consistent with the findings of the CREATE-X trial^4^, patients receiving capecitabine had a better 5 years RFS (66% vs 41%, capecitabine vs no capecitabine, respectively; Hazard Ratio (HR) = 0.47; 95% Confidence Interval (CI) = 0.23 to 0.96; *p =* 0.02). Overall survival (OS) was also better, but the difference was not statistically significant (64% vs 48%, capecitabine vs no capecitabine, respectively; HR = 0.51; 95% CI = 0.22 to 1.14; *p =* 0.072) (**Supplementary Figure 2**).

**Table 1.** Clinical characteristics of TRICIA patients.

| Clinical Characteristics | All Patients (N=93) |
| --- | --- |
| Follow Up from Baseline, Median (Years) [Range] | 3.45 [1.02 - 19.70] |
| Age, Median (Range) | 55.4 (28.0 - 79.1) |
| Stage, n (%) |  |
| 1a | 5 (5) |
| 1b | 2 (2) |
| 2a | 34 (37) |
| 2b | 19 (20) |
| 3a | 18 (19) |
| 3b | 9 (10) |
| 3c | 6 (7) |
| Tumor Size, n (%) |  |
| T0 | 2 (2) |
| T1 | 9 (10) |
| T2 | 50 (54) |
| T3 | 21 (22) |
| T4 | 11 (12) |
| Nodal Status, n (%) |  |
| N0 | 42 (45) |
| N1 | 38 (41) |
| N2 | 7 (8) |
| N3 | 5 (5) |
| Missing | 1 (1) |
| Neoadjuvant Treatment, n (%) |  |
| Taxol | 88 (96) |
| Carboplatin | 38 (41) |
| AC | 80 (87) |
| Atezo/Placebo | 9 (10) |
| Other | 9 (10) |
| Pathological Outcome, n (%) |  |
| RCB 1 | 21 (23) |
| RCB 2 | 46 (50) |
| RCB 3 | 25 (27) |
| Missing | 1 (1) |
| Adjuvant Capecitabine, n (%) |  |
| Yes | 61 (66) |
| Recurrences, n (%) |  |
| Yes | 37 (40) |

Tumor informed personalized ctDNA assays were developed in house with five variants per tumor for a total of 467 variants (**Supplementary Table 1).** Each assay was first validated in the matched patient tumor (**Supplementary Figure 3**) and then used to analyze 262 plasma samples (**Supplementary Table 2**). We controlled for clonal hematopoiesis by using patient’s normal control sample, only 3 variants were identified as clonal hematopoiesis mutation and replaced by additional tumor-specific variants. In total we selected 430 genes and 92% were unique to each patient; only TP53 (n = 20) and PIK3CA (n = 4) were selected in more than 2 patients (**Supplementary Table 2)**. ctDNA was defined as positive if at least one variant was detectable at each time point tested. The proportion of the number of variants detected per patient at each time point is shown in **Supplementary Figure 4**. **Figure 2** shows a patient summary throughout clinical follow-up describing the ctDNA detectability at each time point and patient status.

**Figure 2.**
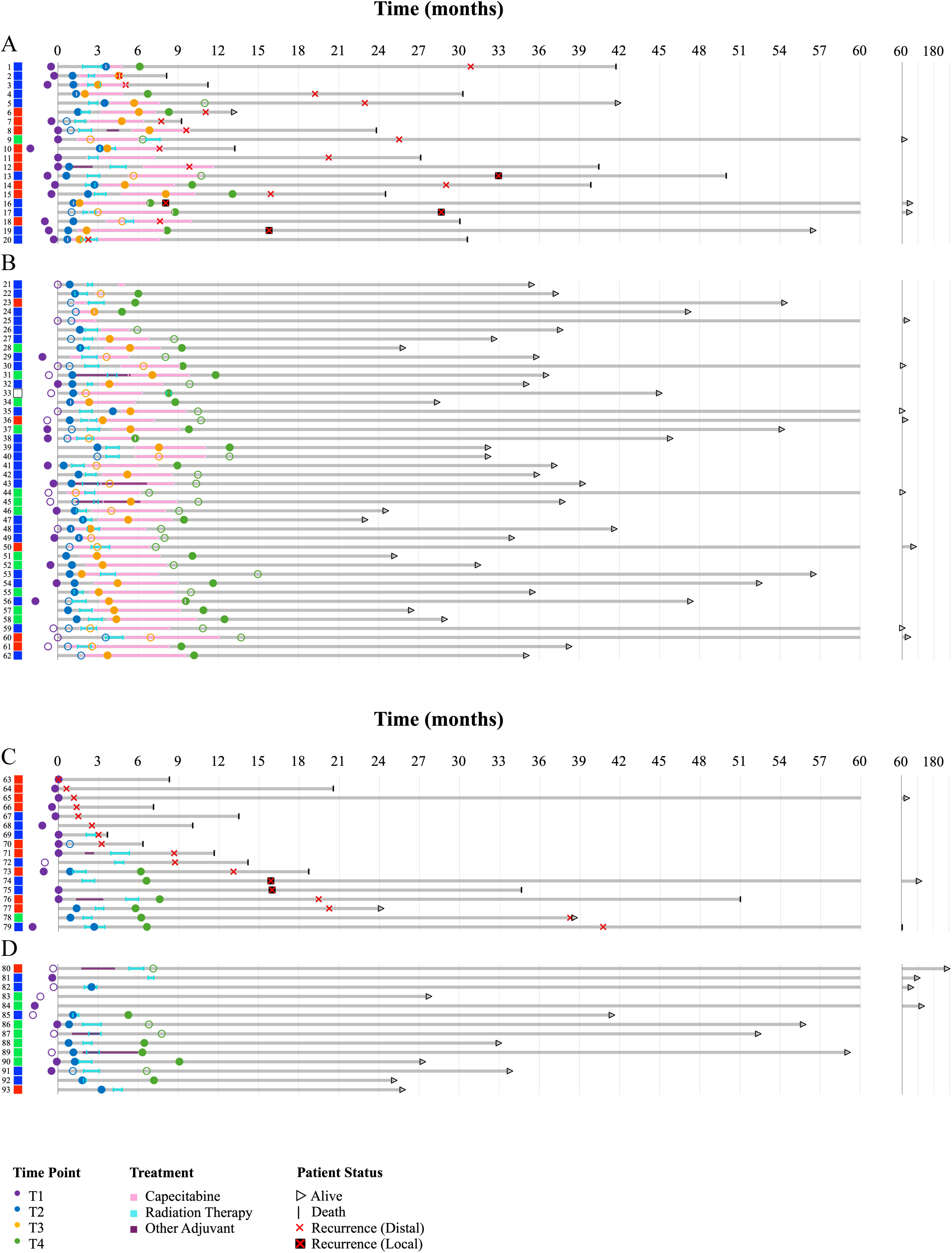
Patient Summary. Swim plot of patients in the TRICIA study. Panel **(A)** shows patients who received capecitabine and relapsed, panel **(B)** shows patients who received capecitabine and who did not relapse. Panel **(C)** shows patients who did not receive capecitabine and relapsed, and panel **(D)** shows patients who did not receive capecitabine and who did not relapse. n = 93.

### Validation of pre-operative ctDNA detection as a strong predictor of patient outcomes

We initially assessed the ctDNA status at the end of NAC and prior to surgery in order to provide an independent validation of the strong prognostic value of ctDNA detection using our assays at this time point, as previously reported by our group^11^. We analyzed T1 samples from 64 patients. Among these, 44 patients were classified as ctDNA-positive (69%), and 20 patients were classified as ctDNA-negative (31%). During the follow-up period, 29 patients experienced disease relapse: 3 had local relapse, while 26 had distant relapses. The number of detectable variants per patient at T1 varied according to patient outcome, no variants were detectable in 54% of non-relapse patients compared to 3% of relapsed patients, and 1 – 2 variants were detectable in 40% of non-relapse patients compared to 55% of relapsed patients.

The overall recurrence rate in ctDNA-positive patients was 64% (28/44) and 5% (1/20) in ctDNA-negative patients at T1 (*p* < 0.001) (**Figure 1B**). This means a negative predictive value (NPV) for no relapse of 95% and a positive predictive value for relapse (PPV) of 64%. These predictive values closely align with those reported in our previous study^11^, where we observed an NPV of 89% and a PPV of 71%. A T1 positive ctDNA test was detected in 28 out of 29 patients who relapsed, yielding a high sensitivity of 97%. In contrast, 19 out of 35 patients who did not relapse tested ctDNA-negative, resulting in 54% specificity.

The RFS for ctDNA-patients was significantly longer than in ctDNA+ patients (median = 55.7 vs 19.5 months, respectively, *p* < 0.0001), with a HR of 0.052 (95% CI = 0.024 – 0.109), reflecting a lower likelihood of relapse **(Figure 1B).** Similarly, OS for ctDNA-patients was markedly improved (*p* = 0.0001), with a HR of 0.06 (95% CI = 0.027 – 0.131), further indicating the favorable prognosis in this cohort **(Figure 1C).** In fact, the 5-year overall survival of the ctDNA-cohort was 95% compared to 35% for the ctDNA+ cohort. The consistency between the findings of this independent cohort and our earlier work further validates the prognostic value of ctDNA at this pre-surgery timepoint as measured by our approach.

We repeated the analysis only on patients receiving adjuvant capecitabine (n = 39). As with the results for the entire cohort, the T1 time point was highly prognostic for RFS (*p* = 0.0009) and OS (*p* = 0.001) **(Figure 1E, F).** The sensitivity and specificity for relapse detection were 100% and 54% respectively. 38% of patients were ctDNA-and the NPV was 100% while the PPV was 58%. The 5-year overall survival for ctDNA-patients at T1 was 100%.

We further investigated the relationship between ctDNA levels and relapse looking at the fractional abundance (FA) of each variant at the T1 time point. We derived corrected FA (cFA) values by subtracting the FAs in normal pooled plasma from the FA in the patient sample for each variant (**Supplementary Table 3**). Any negative cFAs, i.e. values that were lower than the normal plasma pool threshold, were considered as 0 (undetectable). We then compared the mean cFA values between patients who relapsed and those who did not relapse at the T1 time point. The mean cFA for patients who relapsed was 2.1% (range 0.0017 – 20.9) compared to 0.12% (range 0.0005 – 0.79) in patients who did not relapse (unpaired t-test *p* = 0.0275) (**Figure 1D).** Therefore, higher amounts of ctDNA reflected in higher cFA levels at the T1 time point are associated with a greater likelihood of relapse.

### Pre-operative ctDNA can distinguish poor and good outcome patients within RCB 2 and RCB 3 groups

One of the strongest prognostic markers in non-pCR TNBC patients is Residual Cancer Burden (RCB), classifying patients according to the extent of residual cancer at the time of surgery in both the breast and the lymph nodes following neoadjuvant treatment^13^. Of the 92 patients, 21 (23%) patients were RCB 1 (minimal residual disease), 45 (49%) patients were RCB 2 (moderate residual disease), 25 (27%) were RCB 3 (extensive residual disease), and 1 (1%) patient did not have an RCB score. As expected, and as previously shown in TNBC patients^3, 14^, RCB 1 patients had a higher RFS and OS, followed by RCB 2 and RCB 3 patients (**Supplementary Figure 5B**). Two (10%) RCB 1 patients, 17 (38%), RCB 2 patients, and 18 (72%) RCB 3 patients relapsed (**Supplementary Figure 5A**). Of note, the RCB 1 patients had an estimated 81% 5-year RFS in this cohort, similar to what was previously reported in TNBC patients^3, 14^.

At the pre-operative timepoint (T1), we observed a gradual increase in the proportion of ctDNA+ patients with increasing RCB score (**Supplementary Figure 5C**). We also found that the average ctDNA levels (cFA) increased with RCB score: 0.11 (RCB 1), 0.38 (RCB 2) and 2.85 (RCB 3) (*p* = 0.006) **Supplementary Figure 5D**). These results confirm that ctDNA detection and quantity correlate with residual post-NAC tumor burden in the breast and lymph nodes after NAC and prior to surgery.

We next associated ctDNA detection prior to surgery with patient outcomes (RFS and OS) for each RCB category separately. Of the two RCB 1 patients that relapsed, one had a plasma sample at the T1 time point, which was ctDNA+. ctDNA detection at T1 was not significantly associated with RFS or OS (**Figure 3A and B**). However, for RCB 2 patients, ctDNA detection at T1 was significantly associated with RFS (*p* = 0.04, HR = 0.156 (95% CI = 0.048 to 0.503)) and OS (*p* = 0.032, HR = 0.150 (95% CI = 0.045 to 0.503)) (**Figure 3C and D**). In fact, of 30 RCB 2 patients with T1 samples collected, 12 patients developed disease relapse and 92% (11/12) were ctDNA+ at T1. Nine patients were ctDNA-and only one patient relapsed, whereas 11 of 21 (52%) ctDNA+ patients relapsed. In patients with a RCB 3 score, pre-surgical detection of ctDNA was strongly correlated with both RFS and OS (*p* = 0.0008 and *p* = 0.005, respectively) (**Figure 3E and F**). Notably, 100% of RCB 3 patients who were ctDNA-negative at T1 remained relapse-free (100% specificity) and alive at the time of data cut-off, whereas every RCB 3 patient with detectable ctDNA experienced disease relapse (100% sensitivity). All together, these findings highlight that ctDNA can further stratify RCB 2 and RCB 3 patients into poor and good outcome groups, suggesting that ctDNA detection can add significantly to prognosis above and beyond RCB, especially in patients with moderate to extensive residual disease after neoadjuvant chemotherapy. For this reason, we pooled data from RCB 2 and RCB 3 patients together and reanalysed the prognostic value of pre-surgical ctDNA. The absence of ctDNA detection at the T1 time point was associated with a 5-year RFS of 92%, while the detection of ctDNA was associated with a 5-year RFS of 25% (*p* = 0.0002, **Figure 3G**). ctDNA+ patients had an extremely poor 5-year OS with only 10% patients alive compared to 90% for ctDNA-patients (*p* = 0.0003) **(Figure 3H).** In this combined group, the test demonstrated strong performance characteristics, with a positive predictive value (PPV) of 73%, negative predictive value (NPV) of 92%, and sensitivity and specificity of 96% and 55%, respectively. These findings support the use of pre-surgical ctDNA as a unified prognostic test across both RCB 2 and RCB 3 patients to identify low risk patients who may be spared further adjuvant treatment.

**Figure 3.**
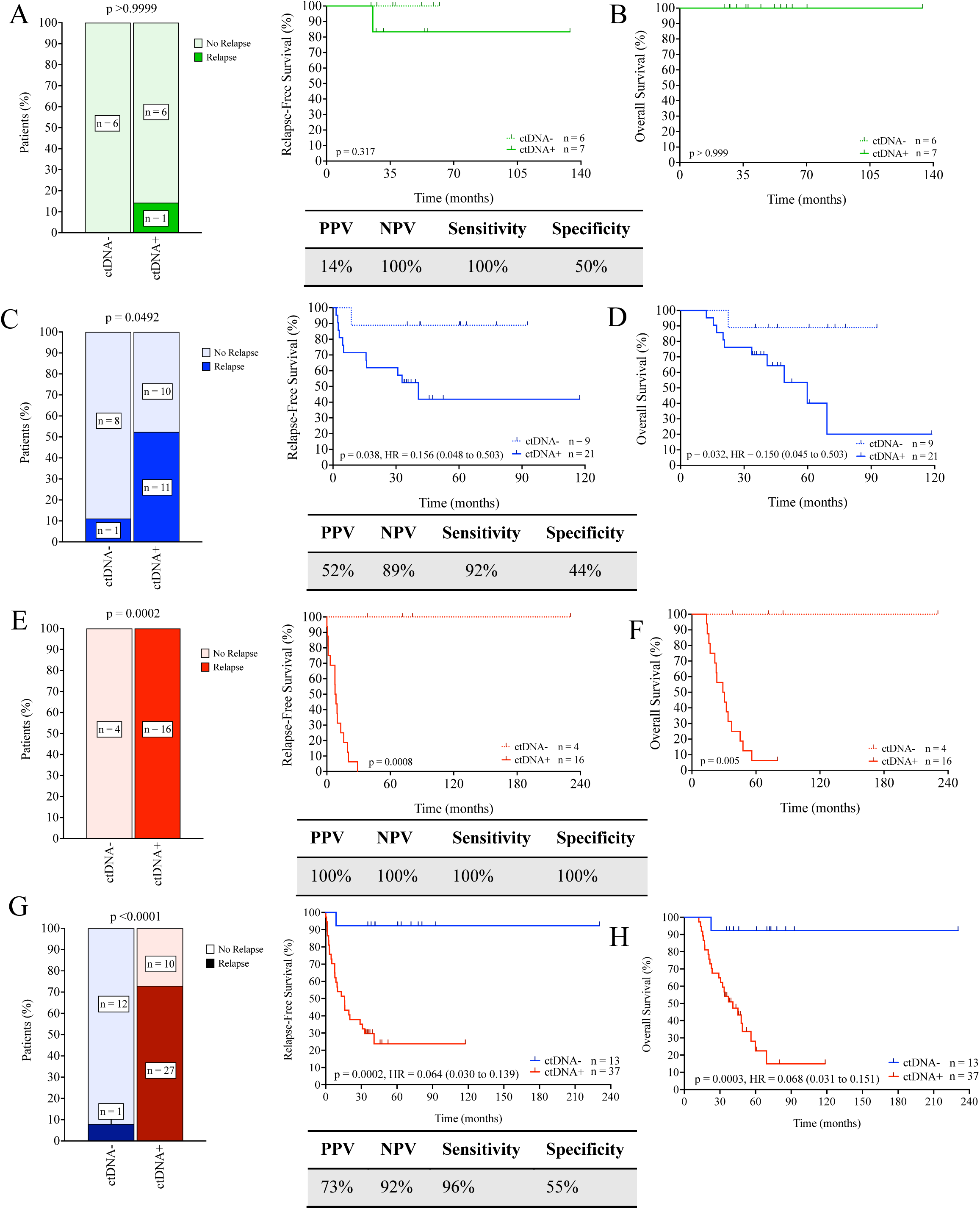
Prognostic performance of ctDNA detection at the post-neoadjuvant, pre-surgical time point (T1) by Residual Cancer Burden Score. Proportions of patients with detectable (ctDNA+) and undetectable (ctDNA-) ctDNA at T1 for **(A)** RCB 1 (n=13), **(C)** RCB 2 (n=30), **(E)** RCB 3 (n=20), and **(G)** RCB 2 and 3 combined (n=50) patients who experienced relapse or remained relapse-free. Statistical comparison of proportions was performed using Fisher’s exact test. Diagnostic performance tables include the PPV, NPV, sensitivity, and specificity of ctDNA status at T1 for predicting relapse. RFS was assessed by Kaplan–Meier (KM) analysis, stratified by ctDNA status. KM analysis of OS stratified by T1 ctDNA status for **(B)** RCB 1 (n=13), **(D)** RCB 2 (n=30), **(F)** RCB 3 (n=20), and **(H)** RCB 2 and 3 combined (n=50). P-values were calculated using the log-rank test and associated hazard ratios.

### CtDNA dynamics and clearance during surgery

As ctDNA can reflect tumor burden changes after surgical intervention, we examined the ctDNA dynamic changes from pre-op (T1) to early post-op (T2) samples. Among the 39 patients with plasma collected at both T1 and T2 time points prior to any adjuvant treatment, 26 were ctDNA+ at T1. Of these, 8 patients (31%) became ctDNA negative after surgery. On the other hand, 7 patients who were initially ctDNA-at T1 became ctDNA+ at T2 **(Supplementary Figure 6A)**. The log rank test showed statistically significant changes in RFS among the different ctDNA detection groups (*p* = 0.0207) **(Supplementary Figure 6B).** Patients with no ctDNA detectable at T1/T2 (-/-) had 100% RFS at our median follow up timepoint of 38 months along with those patients who converted to positive at T2 (-/+) compared to 33% RFS in persistent ctDNA (+/+) patients (*p* = 0.026 and *p* = 0.017, respectively). Cox regression analysis showed that patients who cleared their ctDNA (+/-) following surgery, tended to have better RFS at median follow up (63% relapse rate) than patients who did not clear (+/+) (*p* = 0.47) but poorer RFS than-/-and-/+ patients (*p* = 0.107 and *p* = 0.082, respectively). These data suggest that prognosis was more determined by the presence of ctDNA at the pre-operative time point than by changes in ctDNA occurring during the surgical period and perhaps effected by surgery.

When we looked at the difference in average cFAs between T1 and T2 for these 39 patients we found a dramatic decrease after surgery by 91% (*p* = 0.009) for all variants and by 96% for those variants detectable at T1 (*p* = 0.006) **(Supplementary Figure 6C)**. When we analyzed the different RCB groups, we found drastic decreases in cFA before and after surgery for RCB 2 (96%, *p* = 0.02) and RCB 3 patients (91%, *p* = 0.03), coinciding with the greater tumor burden before surgery in these patients, compared to RCB 1 patients (69%, *p* = 0.234) (**Supplementary Figure 6D)**. Therefore, these results suggest that the extent of ctDNA fall after surgery is proportional to the amount of tumor removed at surgery further supporting ctDNA as a marker of tumor burden.

### ctDNA status during post-operative timepoints predicts relapse and is prognostic

To evaluate MRD detection we analyzed ctDNA detectability in post-operative ctDNA samples for 72 patients at T2, 55 patients at T3 and 66 patients at T4. ctDNA detection at any post-operative timepoint identified 24 out of 26 relapsed patients (sensitivity 92%, specificity 21%) **(Supplementary Figure 7A).** The PPV for relapse detection was 36% with 24 out of 66 ctDNA+ patients experiencing relapse. Among 13 ctDNA-patients, 11 remained relapse-free, resulting in a negative predictive value (NPV) of 85%. Of the two patients who relapsed, ctDNA had been detectable at the T1 time point. For one of these patients, additional samples collected after T4 revealed ctDNA detection 10 months after T4 and 10 months before clinical relapse (**Supplementary Figure 8A**). In fact, the sensitivity of the post-operative test allowed for relapse detection with a median lead time before clinical relapse of 12 months (range 1.57–38.07). The median lead time for local relapses was 15 months (n = 5, range 6.9 – 32.3) versus 12 months (n = 19, range 1.57 – 38.07) for distant relapses, with local relapses having somewhat lower cFA of detectable variants at the time of relapse detection (**Supplementary Figure 8B).**

Patients with no detectable ctDNA at any time point post-op showed a trend toward improved relapse-free survival (RFS 85%) compared to those with detectable ctDNA (58%), with a hazard ratio of 0.34 (95% confidence interval [CI]: 0.13 to 0.92, *p* = 0.130). Similarly, there was a trend toward improved overall survival in ctDNA-patients (RFS 92%) compared to those with detectable ctDNA (51%), with a hazard ratio of 0.202 (95% confidence interval [CI]: 0.066 to 0.618, *p* = 0.079) although these differences were not statistically significant (**Supplementary Figure 7B**).

When analyzing each post-operative time point individually (**Supplementary Figure 7C-F and Figure 4E, F**), ctDNA was detectable in 71% (51/72) of patients at T2, 65% (36/55) at T3 and 59% (39/66) at T4. The rate of relapses tended to be higher in ctDNA+ patients across all time points but was only significant at T4 with 36% of ctDNA+ patients relapsing compared to 11% of ctDNA-patients (*p* = 0.0431). The PPV for relapse detection at the late post-operative T4 timepoint, was 36%, NPV was 89% while sensitivity and specificity were 82% and 49% respectively **(Supplementary Figure 7E)**.

**Figure 4.**
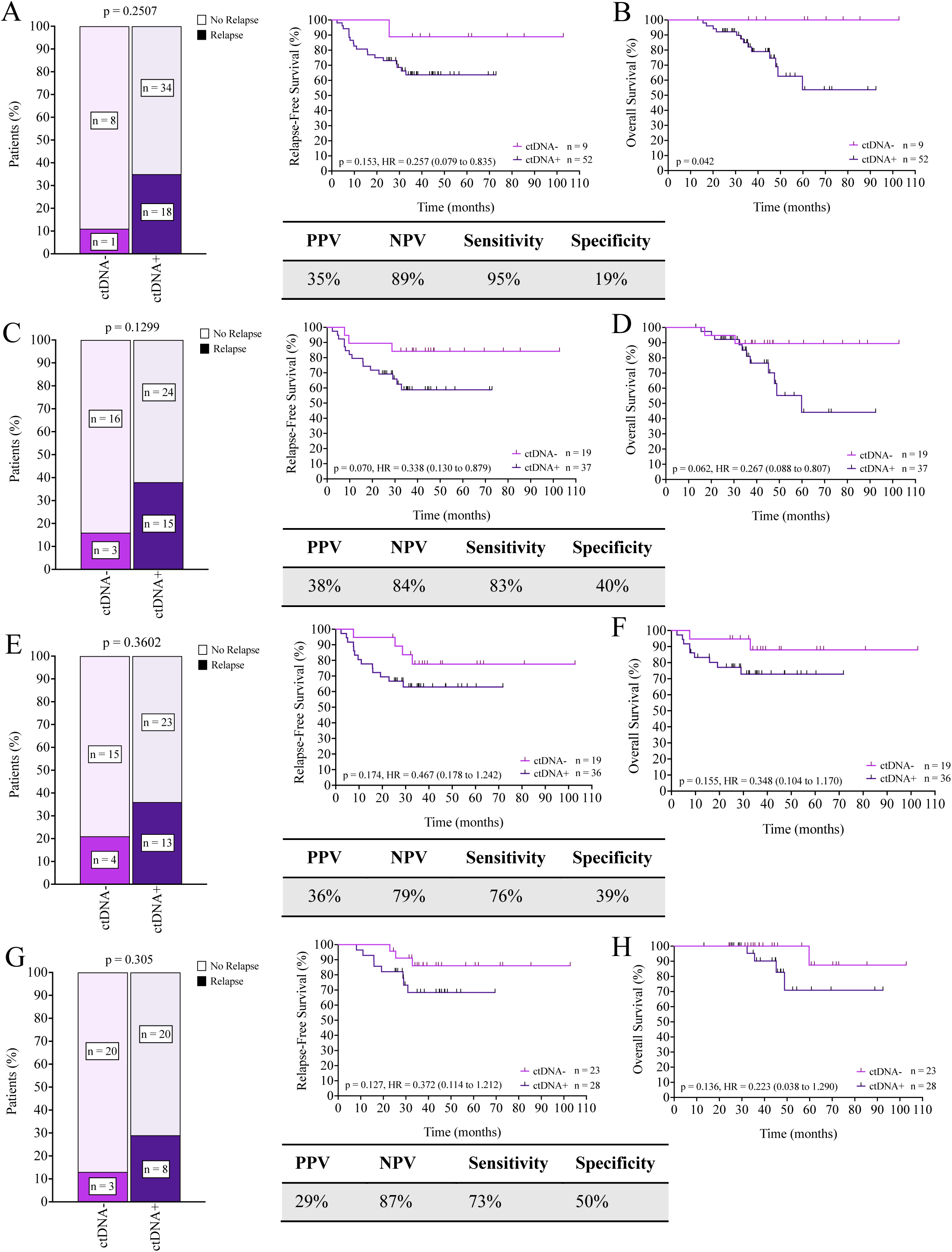
Prognostic performance of ctDNA detection at all post-surgical time points (T2, T3, and T4) for patients who received additional adjuvant capecitabine. Proportions of patients with detectable (ctDNA+) and undetectable (ctDNA-) ctDNA at **(A)** all post-surgical time points (n=61), **(C)** T2 (n=56), **(E)** T3 (n=55), and **(G)** T4 (n=51) in patients who experienced relapse or remained relapse-free. Statistical comparison of proportions was performed using Fisher’s exact test. Diagnostic performance tables include the PPV, NPV, sensitivity, and specificity of ctDNA status at each time point for predicting relapse. RFS was assessed by Kaplan–Meier (KM) analysis, stratified by ctDNA status. KM analysis of OS stratified by ctDNA status for **(B)** all post-surgical time points (n=61), **(D)** T2 (n=56), **(F)** T3 (n=55), and **(H)** T4 (n=51). P-values were calculated using the log-rank test and associated hazard ratios.

The prognosis for patients with undetectable ctDNA (ctDNA-) was generally better than for those with detectable ctDNA (ctDNA+), across all time points, though this difference reached statistical significance only at T4. Specifically, patients with undetectable ctDNA at T4 exhibited significantly longer relapse-free survival (RFS) (*p* = 0.015, HR = 0.24, 95% CI = 0.093 to 0.62) and overall survival (OS) (*p* = 0.036, HR = 0.147, 95% CI = 0.037 to 0.59) **(Supplementary Figure 7E, F)**. Notably, all 27 patients with negative ctDNA at T4 remained alive at the time of our median follow-up of 38 months, and the 5-year OS for ctDNA-patients at T4 was 100%.

We observed that the average concentration of detectable ctDNA (cFA) was significantly higher (> 4-fold) in patients who relapsed compared to those who did not relapse across all post-operative time points (*p* < 0.05), except at T2 (*p* = 0.2) (**Supplementary Figure 8C)**. Indeed, the cFA levels of the relapse patients at this first post-operative time point was much lower than those at T3 and T4, suggesting that ctDNA is being diluted by larger amounts of cfDNA being released as the result of trauma and healing following surgical intervention.

We analyzed separately those patients who received adjuvant capecitabine (**Figure 4A-H**). 67% (39/58) had detectable ctDNA before the initiation of treatment at T2, 65% during treatment at T3 (36/55) and 55% (28/51) after the completion of treatment at T4. Sixty-one patients had at least one post-surgery (T2, T3, and/or T4) collection available for MRD analysis, fifty-two of these patients were ctDNA+ while 9 remained ctDNA negative throughout the adjuvant period (**Figure 4A**). When considering all post-op time points together in patients who received capecitabine, 95% of relapsed patients were ctDNA+ (18/19) and the median lead time to the detection of relapse was 9.3 months (range = 1.6 – 32.3). For those patients who received capecitabine and were ctDNA-showed a trend toward improved RFS (*p* = 0.15) and had significantly higher overall survival than ctDNA+ patients (*p* = 0.042) **(Figure 4A-B).** Survival was 100% at 5 years for ctDNA-patients compared to 54% for ctDNA+ patients. No significant association with RFS or OS was found when we analyzed each time point separately **(Figure 4C-H)**.

### ctDNA dynamics and clearance during the adjuvant period

There were 59 patients with plasma collected at both T2 and T4 time points. 42 patients were ctDNA+ at T2 and, of these, 14 patients (33%) became ctDNA negative at the T4 collection. On the other hand, there were 9 patients that were ctDNA-at T2 that became ctDNA+ at T4 and only one of these patients relapsed (**Supplementary Figure 9A**). In fact, these patients had similar RFS to patients who cleared (+/-) their ctDNA (**Supplementary Figure 9B**). Those patients with persistent ctDNA (+/+) had significantly worse 5-year RFS (51%) than patients with consistently undetectable ctDNA (-/-) (*p* = 0.036) (**Supplementary Figure 9B**). The T4 cFA for detectable variants in the-/+ patients was markedly lower (almost 5-fold lower, *p* = 0.016) than the cFA of detectable variants in the 28 patients with persistent ctDNA (+/+) (**Supplementary Figure 9C**). We therefore looked at whether cFA changes between T2 and T4 were different in relapsed vs non-relapsed patients. We found that patients who relapsed had significantly increased cFA at T4 for all variants tested (average = 0.72) compared to T2 (average = 0.10) (*p* = 0.002) while non-relapsed patients had similar cFA at both time points (0.05 at T2 and 0.06 at T4) (*p* = 0.67) **(Supplementary Figure 9D).** The rising ctDNA signal observed in patients who relapsed indicates an increasing MRD burden and highlights the potential benefit of early therapeutic intervention in these patients, at the end of adjuvant therapy.

We analyzed ctDNA dynamics in the 42 patients with ctDNA+ at T2 that also had blood collected at T4, 32 of these patients received capecitabine while 10 did not. In the 10 non-capecitabine patients, only 1 (9%) cleared and that patient did not show relapse. Of 32 patients receiving adjuvant capecitabine, 13 cleared their ctDNA by the T4 time point (41%), significantly more than in the cohort without capecitabine (*p* < 0.0001) (**Figure 5A, E, F**). Three of these 13 patients with ctDNA clearance had become negative by the T3 time point, while the remainder still had detectable ctDNA at T3, suggesting a continued effect of capecitabine over the 6-month treatment period in the majority of these patients. Two of the 13 patients with clearance showed disease relapse (15%) while the rate of relapse for patients without clearance was 37% (*p* = 0.10) **(Figure 5B).** The relapse-free survival for patients on capecitabine who cleared their ctDNA at T4 tended to be better (83%) than that for those who did not clear (56%), HR of 0.29 (95% CI = 0.079 to 1.08, *p* = 0.10) (**Figure 5B**). The overall survival at 5 years was 100% and 52% in patients who cleared and not cleared respectively but it did not reach statistical significance (*p* = 0.20, HR = 0.26, 95% CI = 0.046 to 1.53) (**Figure 5B)**.

**Figure 5.**
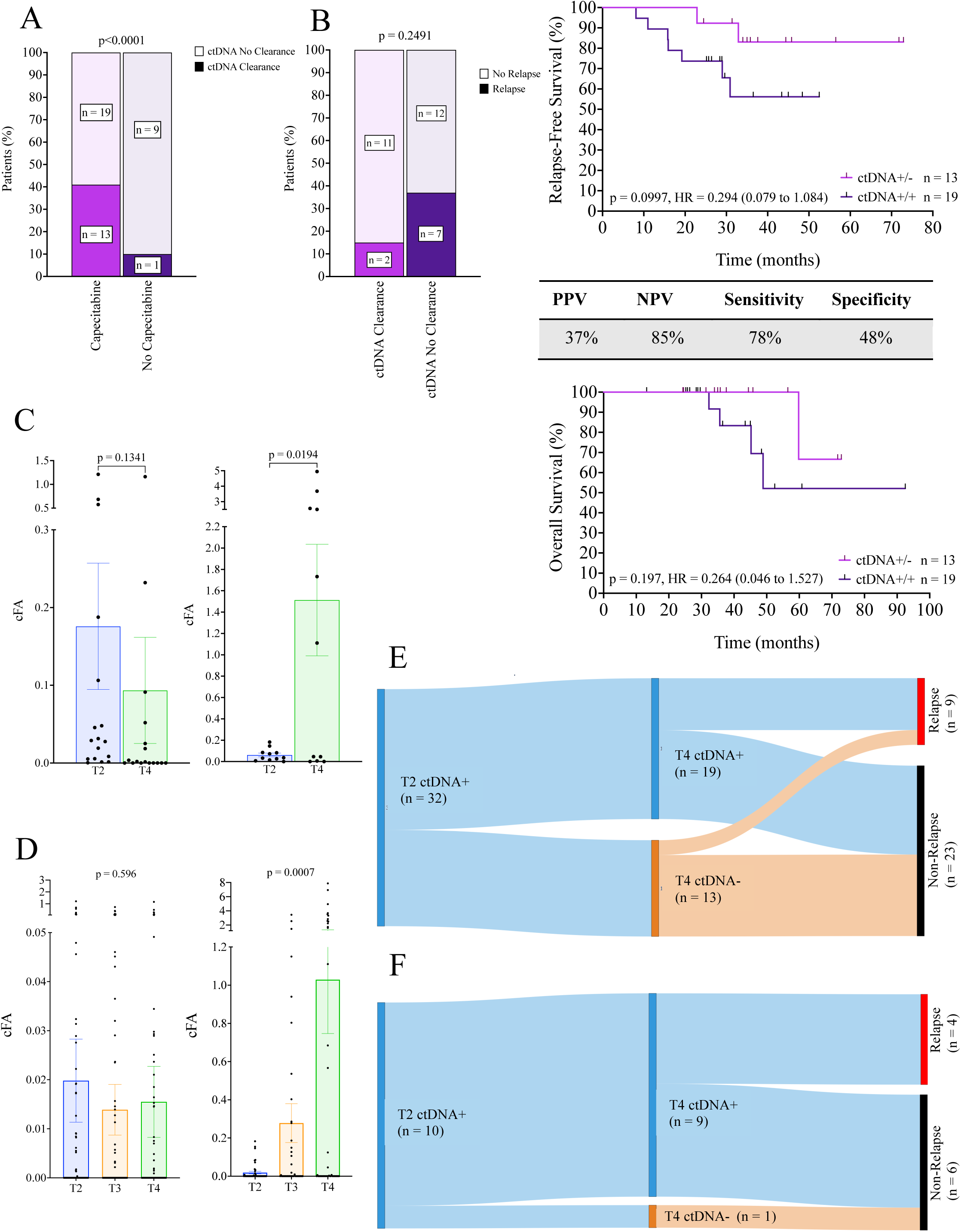
Prognostic performance of changes in ctDNA detection from the first post-surgical time point (T2) to the last post-surgical time point (T4) in patients who received additional adjuvant capecitabine. **(A)** Proportions of patients who received additional adjuvant capecitabine (n=32) and did not receive additional adjuvant capecitabine (n=10), who experienced ctDNA clearance or not. Statistical comparison of proportions was performed using Fisher’s exact test. **(B)** Proportions of patients who received additional adjuvant capecitabine with consistently detectable ctDNA (ctDNA+/+, n=19), or exhibiting ctDNA clearance (ctDNA+/-, n=13) from T2 to T4 who experienced relapse or remained relapse-free. Statistical comparison of proportions was performed using Fisher’s exact test. Diagnostic performance table includes the PPV, NPV, sensitivity, and specificity of ctDNA status at each time point for predicting relapse. RFS and OS were assessed by KM analysis, stratified by changes in ctDNA status. P-value was calculated using the log-rank test and associated hazard ratios. **(C)** Changes in mean cFA (± standard error of the mean, SEM) from T2 to T4 in patients without ctDNA clearance (n=19). Left: analysis of changes in cFA from T2 to T4 of variants detectable at T2 in non-relapsed patients (n=7). Right: analysis of changes in cFA from T2 to T4 of variants detectable at T2 in relapsed patients (n=12). P-values were calculated using a paired two-tailed Student’s t-test. **(D)** Changes in mean cFA (± SEM) at T2, T3, and T4 across all tested variants in patients who received additional adjuvant capecitabine (n=45). Left: analysis restricted to non-relapsed patients (n=36). Right: analysis restricted to relapsed patients (n=9). P-values were calculated using a paired two-tailed Student’s t-test. **(E)** Sankey plot showing dynamic ctDNA status from T2 to T4 in patients who received additional adjuvant capecitabine and were ctDNA+ at T2 (n=32). **(F)** Sankey plot showing dynamic ctDNA status from T2 to T4 in patients who did not receive additional adjuvant capecitabine and were ctDNA+ at T2 (n=10).

Among the 19 patients treated with adjuvant capecitabine who did not clear their ctDNA, those who remained relapse-free (n = 12) exhibited a trend toward decreased cFA from T2 to T4 (*p* = 0.13), whereas those who experienced relapse (n = 7) showed a significant increase in cFA (*p* = 0.019) **(Figure 5C).** In these 7 relapsed patients, the cFA continuously increased by 93% from T2 to T3 and by 73% from T3 to T4 (*p* = 0.0007), unlike in patients who did not relapse (**Figure 5D**). Therefore, our results demonstrate that there is increased ctDNA clearance in patients treated with capecitabine and that MRD tumor burden tends to decline, even in the absence of complete ctDNA clearance, in patients who remain relapse-free.

## Discussion

CtDNA has been used as a plasma-based biomarker to monitor treatment and tumor progression in many cancers, including breast cancer^15–19^. The detection of ctDNA using tumor-informed assays has been shown to be highly sensitive for the detection of minimal residual disease in breast cancer^18, 20–22^. The present study is the largest reported series of TNBC patients with residual disease at surgery with serial ctDNA measurements before surgery (T1), early after surgery (T2), during (T3) and immediately after capecitabine treatment (T4). Using a unique approach to tumor-informed ctDNA detection in 92 non-pCR TNBC patients, we found that our ctDNA test allows the stratification of prognostic groups in the neoadjuvant and adjuvant setting for the management of non-pCR TNBC patients. We were able to predict 97% of relapses at a median of 12 months prior to clinical relapse in this poor risk group of TNBC patients. The very high negative predictive value and sensitivity of our test allowed us to distinguish patients with excellent prognosis from those with poor prognosis. We observed that our test behaves with different sensitivity and specificity at each time point. Of the four time points tested, ctDNA detection at the post-NAC pre-surgery time point (T1) clearly had the strongest prognostic value for both RFS and OS, reaching an RFS of 95% and an OS of 95%. These results validate our previously reported results in an independent cohort of high-risk TNBC patients^11^. Indeed only 1 ctDNA-patient (3%) at this time point showed distant disease relapse, suggesting that the lack of ctDNA detectability at this time point has sufficient prognostic power to consider proposing trials of adjuvant therapy de-escalation.

These results are among the best reported ctDNA MRD results using a single landmark time point (T1) and highlight the validity of our tumor informed 5-variant pre-amplification ddPCR approach, which can be performed in hospital laboratories. Interestingly, the corrected variant fraction abundances at the pre-operative time point (T1) reflected the amount of residual disease found at surgery. Moreover, in patients who eventually developed relapses, all 3 of the local relapses with the T1 time point were also ctDNA+ at T1, suggesting that this test at T1 can predict local relapses as well as distant relapses.

In the MRD setting (once the tumor has been removed), sensitivity and specificity was less than at the T1 time point, and the best of these post-operative time points is the T4 late post-op/after capecitabine time point. The weaker prognostic effect of the first post-operative time point (T2) may be due to post-surgical dilution of ctDNA by cell free DNA originating from traumatized and repairing tissues as previously described^23^. In fact, it has been shown that there is an influx of trauma-induced cfDNA release following surgery that can persist up to 4 weeks^23^. These studies suggest a secondary plasma sample be taken after 4 weeks for patients who are initially post-operatively ctDNA-. Given the pre-amplification procedure in our protocol, we could not measure the absolute quantity of cell free DNA in our samples. However, the amount of ctDNA (cFA) detected was several folds lower at T2 compared to the other time points in patients who relapsed. In fact, 3 of the 4 patients who were ctDNA-at T2 and relapsed were later ctDNA+ prior to relapse, and all these patients had their plasma sample taken within that 4-week period following surgery.

The extent of residual disease is an important prognostic factor in TNBC patients receiving neoadjuvant therapy. Previous studies have demonstrated an association between ctDNA detection and RCB score^24,25^. We found that ctDNA can distinguish good from poor prognosis very well especially in patients with more extensive residual disease, RCB 2 and RCB 3 patients. While RCB 1 patients had the best outcomes regardless of ctDNA status at T1, RCB 2 and RCB 3 patients showed a significant association between ctDNA positivity at T1 and worse RFS and OS. Notably, all RCB 3 patients who were ctDNA-at T1 remained relapse-free, whereas those who were ctDNA+ all experienced relapse, highlighting the potential of our ctDNA test to further stratify relapse risk within the RCB 3 subgroup with 100% sensitivity and 100% specificity. These findings suggest that ctDNA detection may be superior or complimentary to RCB scoring in assessing prognosis in non-pCR TNBC patients, indeed ctDNA may be more clinically useful in the RCB 2 – RCB 3 group of patients.

Given the collection of plasma at 4 time points, we were also able to study ctDNA dynamics both pre/post-surgery and pre/post capecitabine therapy. The impact of surgery on ctDNA levels were significant, particularly in patients with the most residual disease (RCB 3), which showed the greatest fall in ctDNA proportions from T1 to T2. This suggests that the magnitude of ctDNA reduction post-surgery reflects the volume of tumor resected, further supporting ctDNA as a dynamic marker of tumor burden. These findings are clinically meaningful as these are the patients most at risk of relapse and thus most likely to receive adjuvant capecitabine. The ability of ctDNA to discriminate bad from poor prognosis in precisely these patients has the potential to enable the greatest impact in chemotherapy reduction. In addition, capecitabine treatment was associated with a significantly higher rate of ctDNA clearance (41% vs 10%), with patients having cleared their ctDNA during the adjuvant period (from T2 to T4) showing excellent prognosis. In contrast, patients who relapse show increases in ctDNA concentration during the adjuvant period. These findings suggest that monitoring ctDNA dynamics during adjuvant therapy may help identify patients who could be candidates for clinical trials exploring therapeutic escalation beyond capecitabine. Interestingly, the mid-adjuvant time point was not adequate to identify all cleared patients, suggesting that 3 months of capecitabine may not be sufficient to prevent relapse. Overall, these findings support ctDNA as a more sensitive indicator of micrometastatic tumor burden than of the primary tumor.

Our approach of mutant variant detection has several challenges. First, all tumor-informed assays for each patient with TNBC are unique and rarely overlap with others, making the generation of these assays labor intensive and expensive. Second, the expected range of mutant allele presence is very low, and detectability is limited by the sensitivity of the ddPCR system, therefore false negatives can occur. In our workflow we have introduced a pre-amplification step enabling the measurement of all five variants to be performed with only 2ml of plasma, which is relatively small quantity for routine blood collections. Although the variant allele frequency is not distorted by the pre-amplification step^12^, we do not rule out the possibility that false positives may be introduced due to polymerase errors, thus lowering the specificity of the test for MRD testing. Despite these limitations, the performance of the post-NAC/pre-operative T1 time point was outstanding in predicting the absence of relapse with specificity >95%. Perhaps because of logistical difficulties in obtaining plasma samples at this time point, the post-NAC pre-op time point has only been studied in a few previous reports. One study sequenced ctDNA using a fixed panel of 275 genes and found that end-of-treatment ctDNA status is prognostic and associates with RCB score, but the performance of their test was not optimal to guide adjuvant therapy^24^. More recently, using a tumor informed method with a 1021 gene panel to guide cfDNA NGS analysis found an association with poor outcome in post-NAC samples but with suboptimal NPV (<90%)^26^. The latest generation “ultra-sensitive” assays in which >1000 genes are probed simultaneously is showing the potential to significantly improve these results^27^. The robustness of our test in the post-NAC T1 time point, as observed now in 2 separate TNBC studies, suggest that this personalized approach to treatment management is ready for prospective testing of its clinical utility in patients who have undergone NAC and require additional adjuvant therapy.

## Supporting information

Summplementary Tables 1-3

## Data Availability

All data produced in the present study are available upon reasonable request to the authors.

## Acknowledgements

We acknowledge funding from CQDM, Exactis Innovation, My Intelligent Machines, and the Jewish General Hospital Foundation. KB is supported by Oncopole, AA-M is supported by the Guerrera Family Cancer Scientist award and the MCTRC Teamsters liquid biopsy program. We would like to thank the Quebec Breast Cancer Foundation and the FRQS Réseau de Recherche Cancer for supporting the biobank infrastructure essential to this study. We thank Lilian Canetti and Naciba Belimane for their expert assistance with pathology services. Sequencing was performed with the support of the IRIC sequencing platform at Université de Montreal and the McGill University Innovation Centre. We thank Kalan Lynn and Nathalie Tremblay for patient recruitment.

## References

1. Yin, L., Duan, J.J., Bian, X.W., et al. (2020). Triple-negative breast cancer molecular subtyping and treatment progress. Breast Cancer Res, 22(1), 61. 10.1186/s13058-020-01296-5

2. Cortazar, P., Zhang, L., Untch, M., et al. (2014). Pathological complete response and long-term clinical benefit in breast cancer: the CTNeoBC pooled analysis. Lancet, 384(9938), 164–172. 10.1016/S0140-6736(13)62422-8

3. Hamy, A.S., Darrigues, L., Laas, E., et al. (2020). Prognostic value of the Residual Cancer Burden index according to breast cancer subtype: Validation on a cohort of BC patients treated by neoadjuvant chemotherapy. PloS one, 15(6), e0234191. 10.1371/journal.pone.0234191

4. Masuda, N., Lee, S.J., Ohtani, S., et al. (2017). Adjuvant capecitabine for Breast Cancer after Preoperative Chemotherapy. N Engl J Med, 376(22), 2147–2159. 10.1056/NEJMoa1612645

5. Dawson, S. J., Tsui, D. W., Murtaza, M., et al. (2013). Analysis of circulating tumor DNA to monitor metastatic breast cancer. N Engl J Med, 368(13), 1199–1209. 10.1056/NEJMoa1213261

6. Khatami, F., & Tavangar, S.M. (2018). Circulating tumor DNA (ctDNA) in the era of personalized cancer therapy. J Diabetes Metab Disord, 17(1), 19–30. 10.1007/s40200-018-0334-x

7. Nicolini, A., Ferrari, P., & Duffy, M.J. (2018). Prognostic and predictive biomarkers in breast cancer: Past, present and future. Semin Cancer Biol, 52(Pt 1), 56–73. 10.1016/j.semcancer.2017.08.010

8. Schwarzenbach, H., Hoon, D.S., & Pantel, K. (2011). Cell-free nucleic acids as biomarkers in cancer patients. Nat Rev Cancer, 11(6), 426–437. 10.1038/nrc3066

9. Arisi, M. F., Dotan, E., & Fernandez, S. V. (2022). Circulating Tumor DNA in Precision Oncology and Its Applications in Colorectal Cancer. International journal of molecular sciences, 23(8), 4441. 10.3390/ijms23084441

10. Garcia-Murillas, I., Schiavon, G., Weigelt, B., et al. (2015). Mutation tracking in circulating tumor DNA predicts relapse in early breast cancer. Sci Transl Med, 7(302), 302ra133. 10.1126/scitranslmed.aab0021

11. Cavallone, L., Aguilar-Mahecha, A., Lafleur, J., et al. (2020). Prognostic and predictive value of circulating tumor DNA during neoadjuvant chemotherapy for triple negative breast cancer. Sci Rep, 10(1), 14704. 10.1038/s41598-020-71236-y

12. Cavallone, L., Aldamry, M., Lafleur, J., et al. (2019). A Study of Pre-Analytical Variables and Optimization of Extraction Method for Circulating Tumor DNA Measurements by Digital Droplet PCR. Cancer Epidemiol Biomarkers Prev, 28(5), 909–916. 10.1158/1055-9965.EPI-18-0586

13. Symmans, W. F., Peintinger, F., Hatzis, C., et al. (2007). Measurement of residual breast cancer burden to predict survival after neoadjuvant chemotherapy. J Clin Oncol, 25(28), 4414–4422. 10.1200/JCO.2007.10.6823

14. Symmans WF, Wei C, Gould R, Yu X, Zhang Y, Liu M, Walls A, Bousamra A, Ramineni M, Sinn B, Hunt K, Buchholz TA, Valero V, Buzdar AU, Yang W, Brewster AM, Moulder S, Pusztai L, Hatzis C, Hortobagyi GN. Long-Term Prognostic Risk After Neoadjuvant Chemotherapy Associated With Residual Cancer Burden and Breast Cancer Subtype. J Clin Oncol. 2017 Apr 1;35(10):1049–1060. doi: 10.1200/JCO.2015.63.1010. Epub 2017 Jan 30. PMID: 28135148; PMCID: PMC5455352.

15. Abbosh C., Birkbak N.J., Wilson G.A., et al. (2017). Phylogenetic ctDNA analysis depicts early-stage lung cancer evolution. Nature, 545, 446–51. 10.1038/nature22364

16. Christensen E., Birkenkamp-Demtröder K., Sethi H., et al. (2019). Early detection of metastatic relapse and monitoring of therapeutic efficacy by Ultra-Deep sequencing of plasma cell-free DNA in patients with urothelial bladder carcinoma. J Clin Oncol, 37, 1547–57. 10.1200/JCO.18.02052

17. Coombes R.C., Armstrong A., Ahmed S. (2019). Early detection of residual breast cancer through a robust, scalable and personalized analysis of circulating tumour DNA (ctDNA) antedates overt metastatic recurrence. Cancer Res, 79.

18. Reinert T., Henriksen T.V., Christensen E., et al. (2019). Analysis of plasma cell-free DNA by ultradeep sequencing in patients with stages I to III colorectal cancer. JAMA Oncol, 5, 1124–10.1001/jamaoncol.2019.0528

19. Tarazona N., Henriksen T.V., Carbonell-Asins J.A., et al. (2020). Circulating tumor DNA to detect minimal residual disease, response to adjuvant therapy, and identify patients at high risk of recurrence in patients with stage I-III CRC. JCO, 38, 4009. 10.1200/JCO.2020.38.15_suppl.4009

20. Natera. [cited 2025 June 5]. Available from: https://www.natera.com/wp-content/uploads/2020/11/Oncology-Clinical-Seeing-beyond-the-limit-Detect-residual-disease-and-assess-treatment-response-SGN_AV_WP.pdf

21. Garcia-Murillas, I., Cutts, R., Abbott, C., et al. (2024). Ultra-sensitive ctDNA mutation tracking to identify molecular residual disease and predict relapse in patients with early breast cancer. JCO 42, 1010–1010(2024). DOI:10.1200/JCO.2024.42.16_suppl.1010

22. Panet, F., Papakonstantinou, A., Borrell, M. et al. Use of ctDNA in early breast cancer: analytical validity and clinical potential. npj Breast Cancer 10, 50 (2024). 10.1038/s41523-024-00653-3

23. Henriksen, T.V., Reinert, T., Christensen, E., et al. (2020). The effect of surgical trauma on circulating free DNA levels in cancer patients-implications for studies of circulating tumor DNA. Molecular oncology, 14(8), 1670–1679. 10.1002/1878-0261.12729

24. Stecklein SR, Kimler BF, Yoder R, Schwensen K, Staley JM, Khan QJ, O’Dea AP, Nye LE, Elia M, Heldstab J, Home T, Hyter S, Isakova K, Pathak HB, Godwin AK, Sharma P. ctDNA and residual cancer burden are prognostic in triple-negative breast cancer patients with residual disease. NPJ Breast Cancer. 2023 Mar 6;9(1):10. doi: 10.1038/s41523-023-00512-7. PMID: 36878909; PMCID: PMC9988835.

25. Parsons HA, Blewett T, Chu X, Sridhar S, Santos K, Xiong K, Abramson VG, Patel A, Cheng J, Brufsky A, Rhoades J, Force J, Liu R, Traina TA, Carey LA, Rimawi MF, Miller KD, Stearns V, Specht J, Falkson C, Burstein HJ, Wolff AC, Winer EP, Tayob N, Krop IE, Makrigiorgos GM, Golub TR, Mayer EL, Adalsteinsson VA. Circulating tumor DNA association with residual cancer burden after neoadjuvant chemotherapy in triple-negative breast cancer in TBCRC 030. Ann Oncol. 2023 Oct;34(10):899–906. doi: 10.1016/j.annonc.2023.08.004. Epub 2023 Aug 18. PMID: 37597579; PMCID: PMC10898256.

26. Li S, Lai H, Liu J, Liu Y, Jin L, Li Y, Liu F, Gong Y, Guan Y, Yi X, Shi Q, Cai Z, Li Q, Li Y, Zhu M, Wang J, Yang Y, Wei W, Yin D, Song E, Liu Q. Circulating Tumor DNA Predicts the Response and Prognosis in Patients With Early Breast Cancer Receiving Neoadjuvant Chemotherapy. JCO Precis Oncol. 2020 Mar 27;4:PO.19.00292. doi: 10.1200/PO.19.00292. PMID: 32923909; PMCID: PMC7450928.

27. Garcia-Murillas I, Abbott CW, Cutts RJ, Boyle SM, Pugh J, Keough KC, Li B, Pyke RM, Navarro FCP, Chen RO, Dunne K, Bunce C, Johnston SRD, Ring A, Russell S, Evans A, Skene A, Smith IE, Turner NC. Whole genome sequencing-powered ctDNA sequencing for breast cancer detection. Ann Oncol. 2025 Feb 4:S0923–7534(25)00053-5. doi: 10.1016/j.annonc.2025.01.021. Epub ahead of print. PMID: 39914664.

