## Supplementary Figures 1-9 for "Clinical Validation of Digital PCR-based ctDNA detection for risk stratification in residual triple negative breast cancer: TRICIA trial results"

### Slide 1
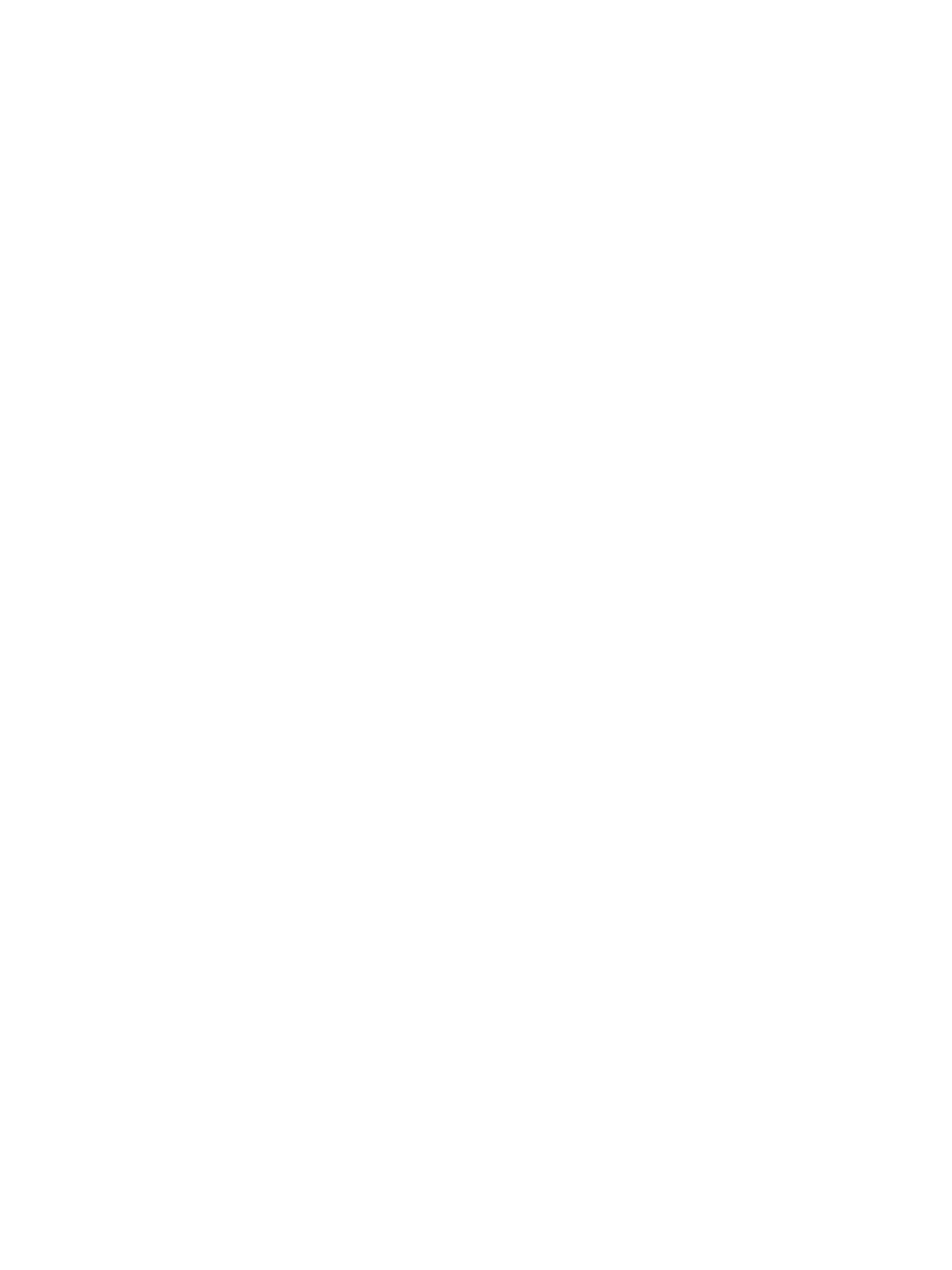

### Slide 2
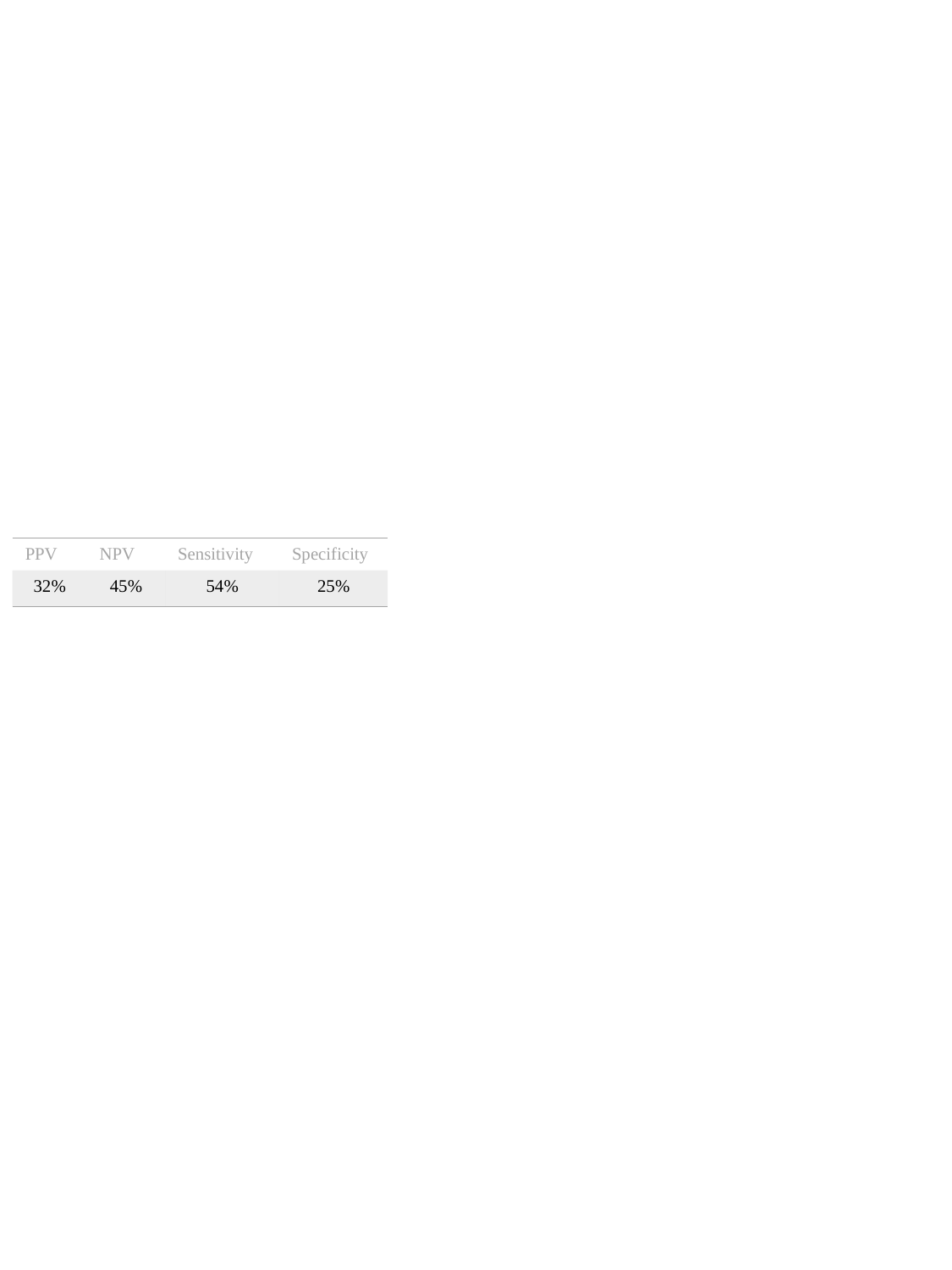

| PPV | NPV | Sensitivity | Specificity |
| --- | --- | --- | --- |
| 32% | 45% | 54% | 25% |

### Slide 3
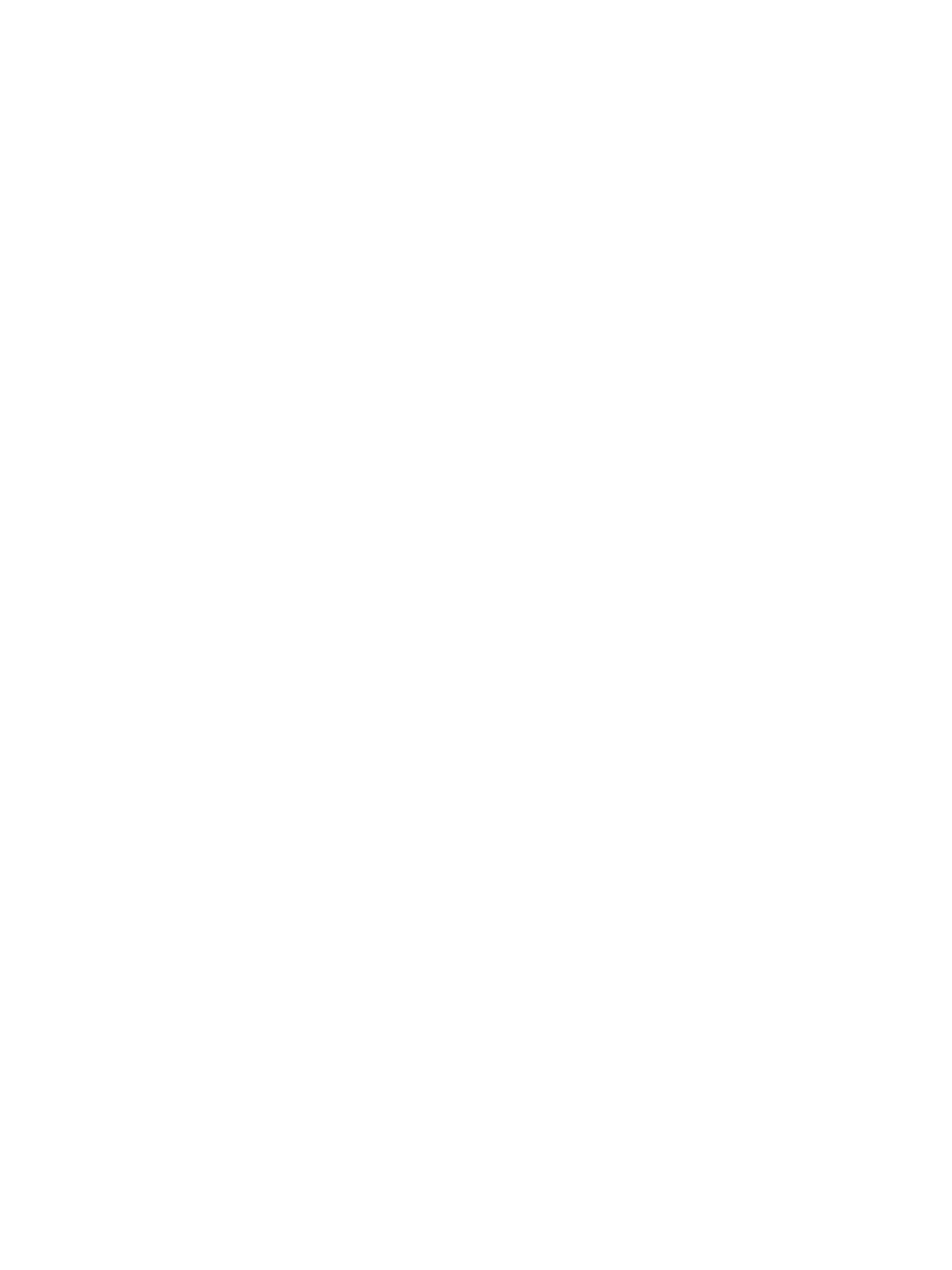

### Slide 4
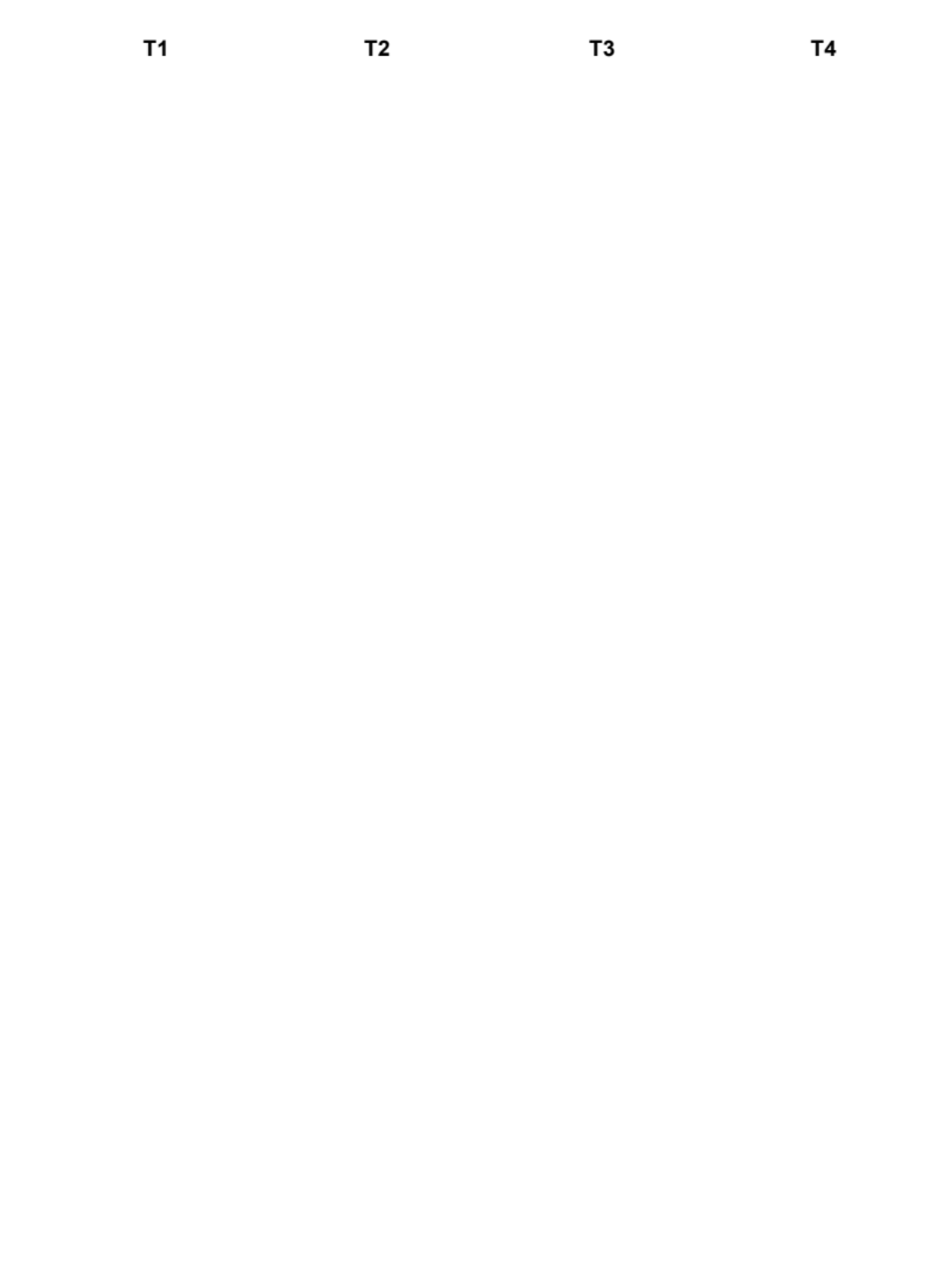

### Slide 5
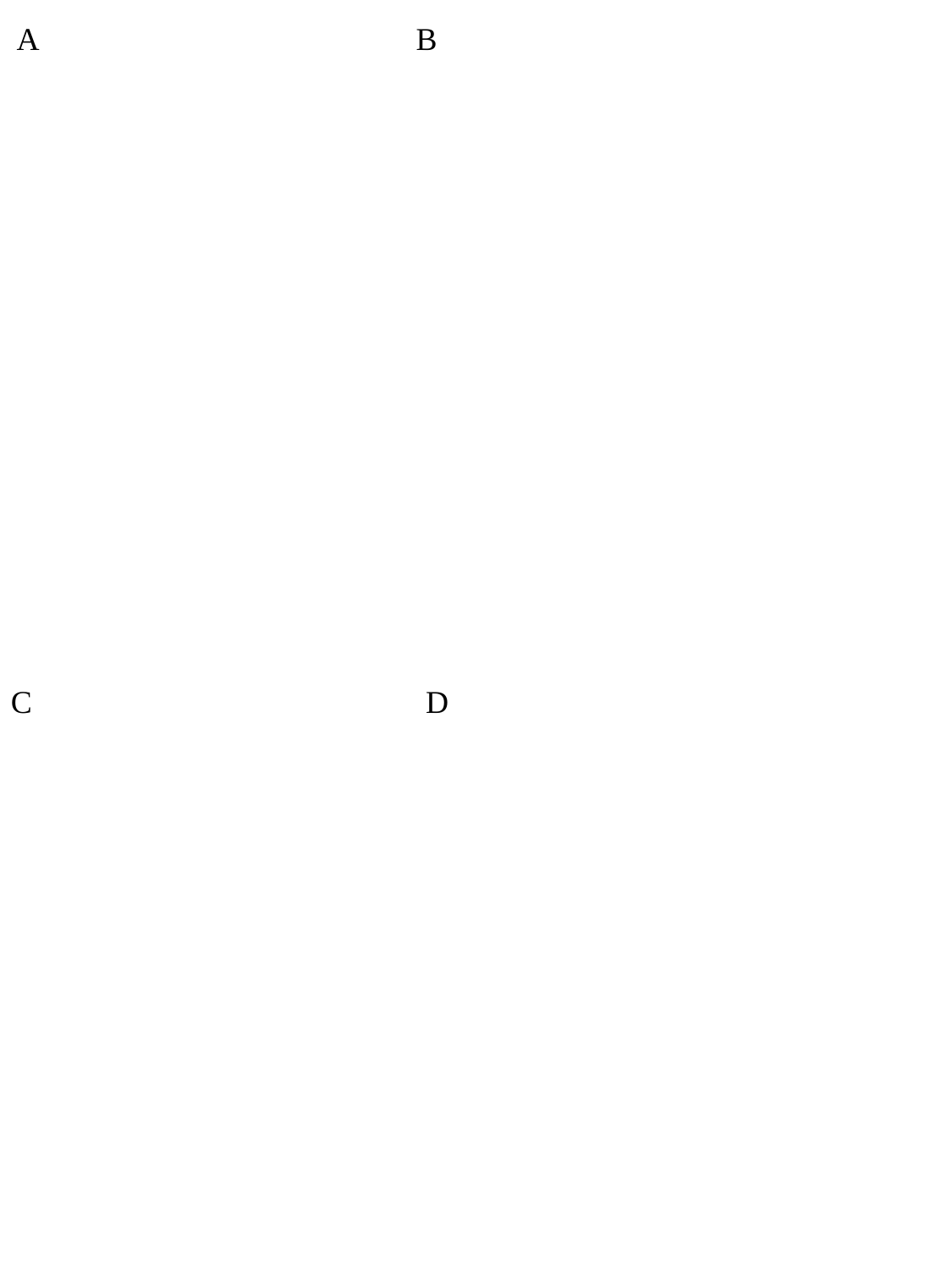

A
B
C
D

### Slide 6
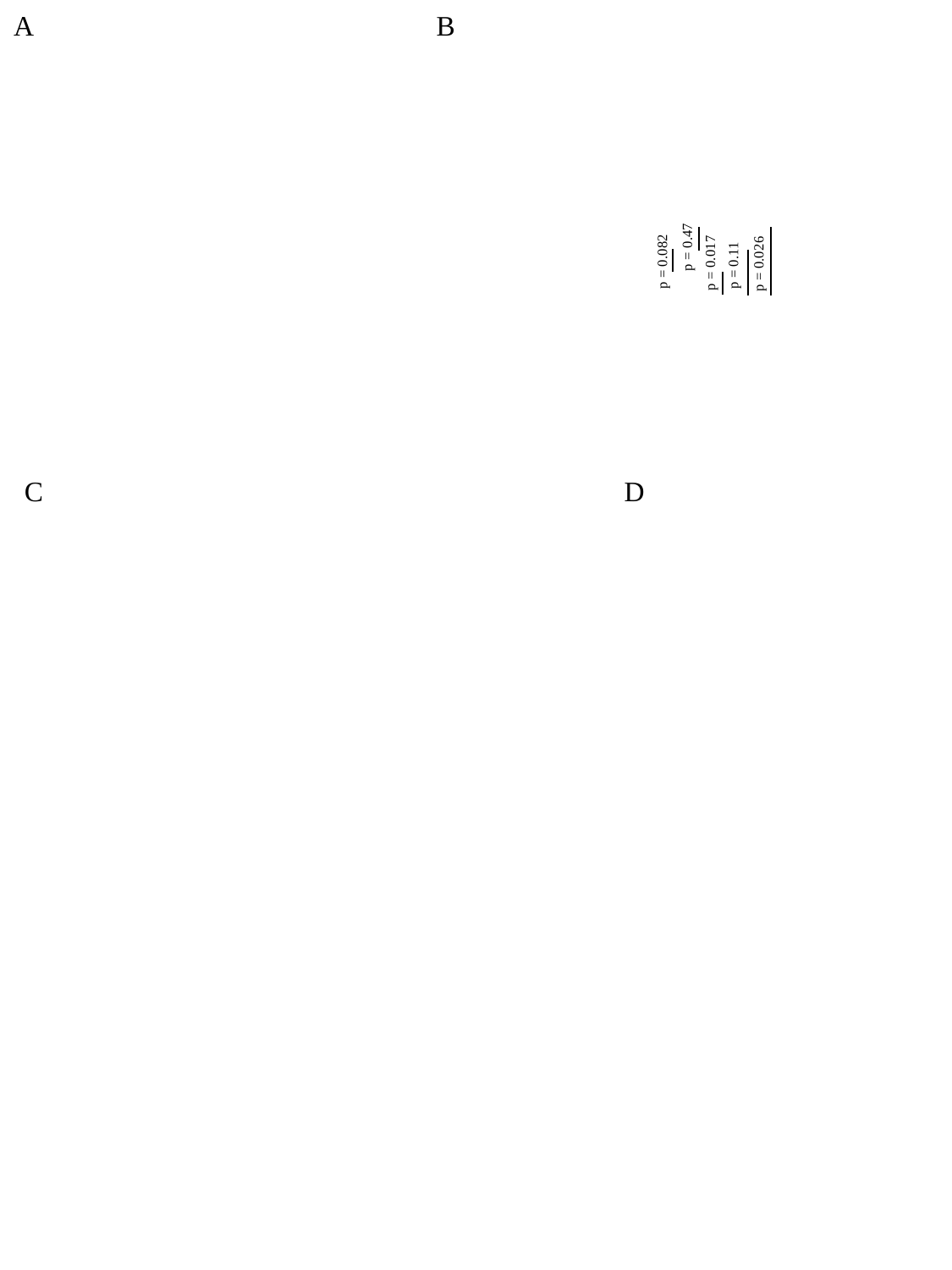

A
B
p = 0.47
p = 0.082
p = 0.017
p = 0.026
p = 0.11
C
D

### Slide 7
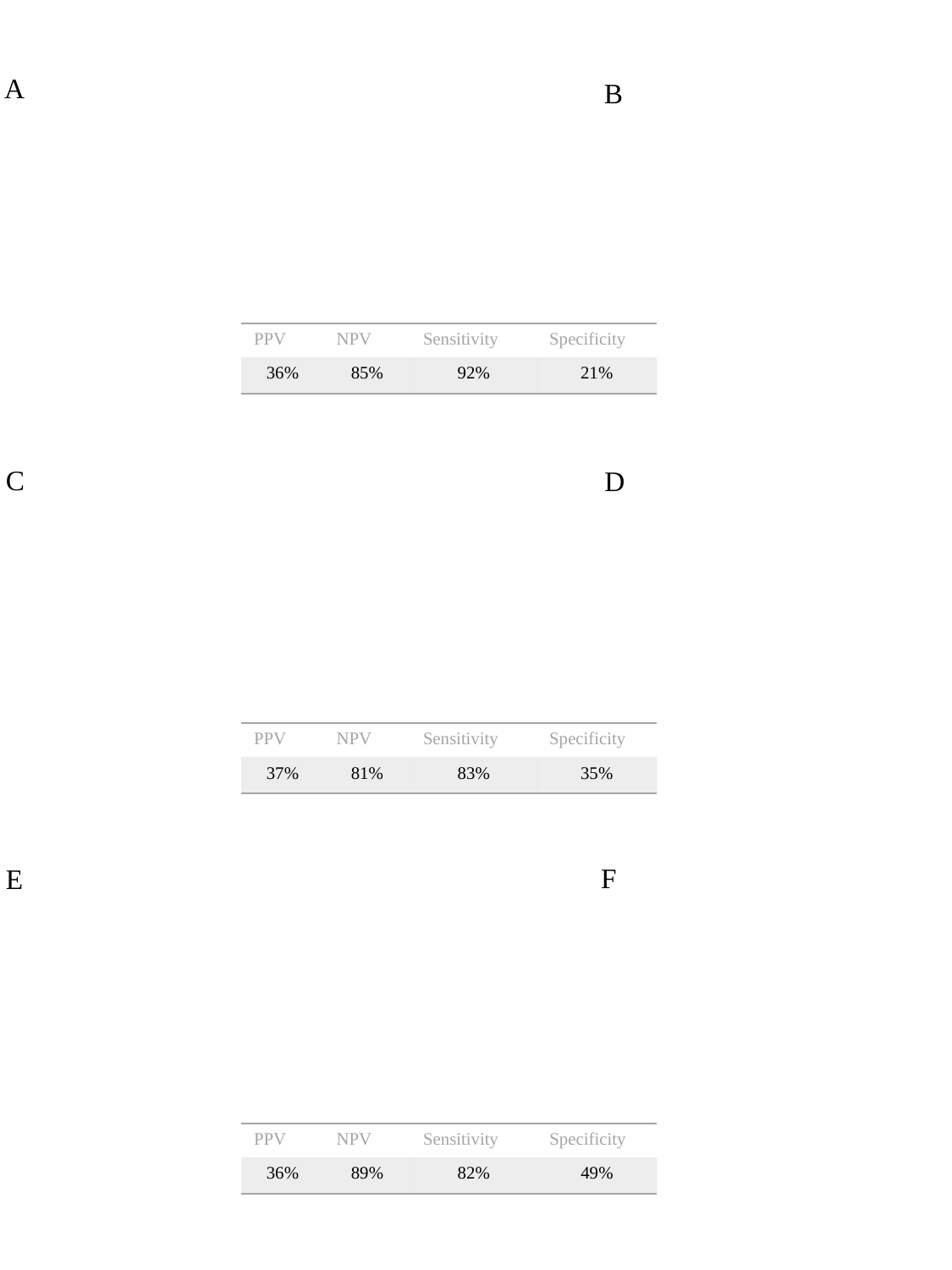

A
B
| PPV | NPV | Sensitivity | Specificity |
| --- | --- | --- | --- |
| 36% | 85% | 92% | 21% |
C
D
| PPV | NPV | Sensitivity | Specificity |
| --- | --- | --- | --- |
| 37% | 81% | 83% | 35% |
F
E
| PPV | NPV | Sensitivity | Specificity |
| --- | --- | --- | --- |
| 36% | 89% | 82% | 49% |

### Slide 8
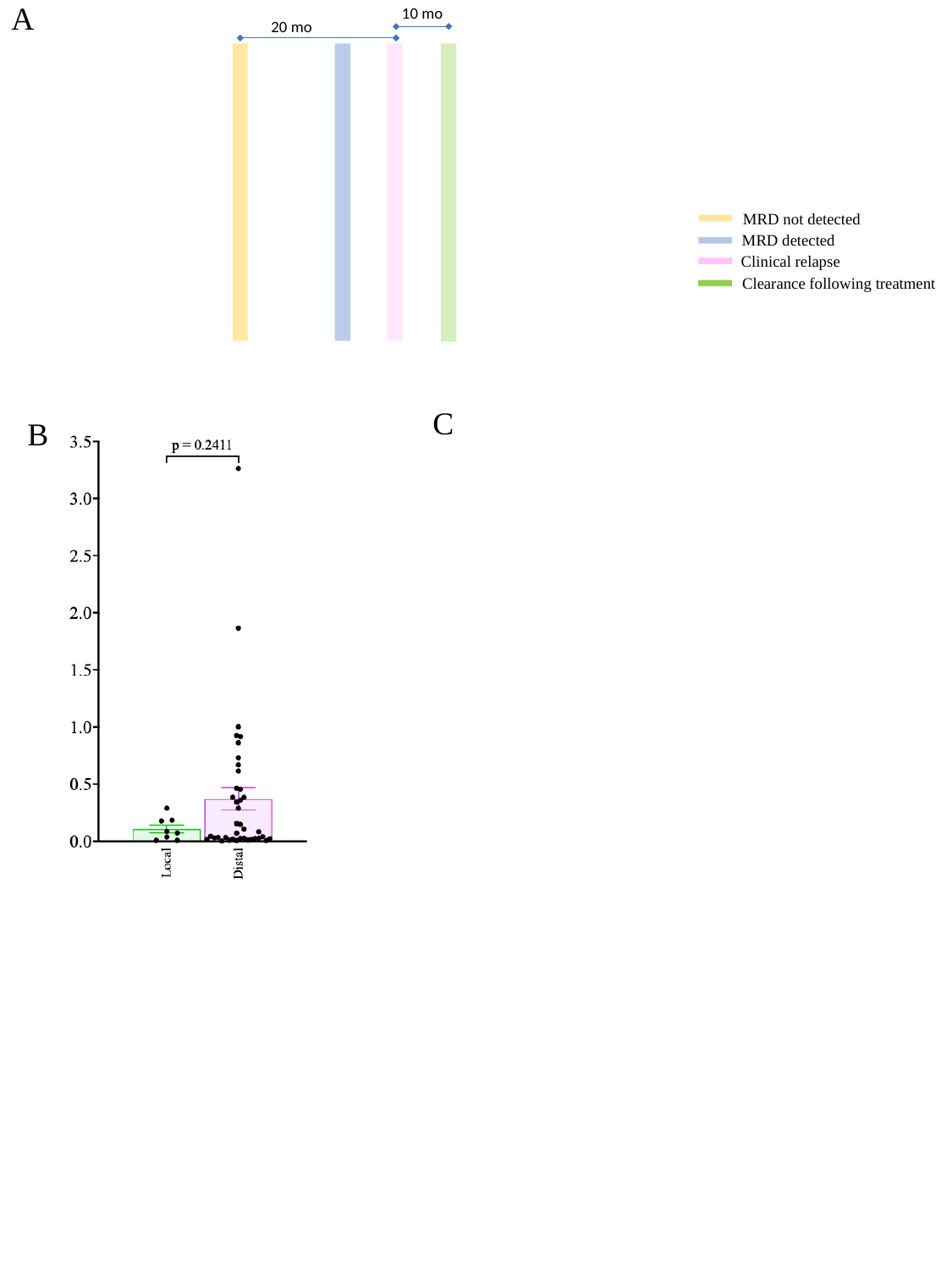

A
10 mo
20 mo
MRD not detected
MRD detected
Clinical relapse
Clearance following treatment
C
B

### Slide 9
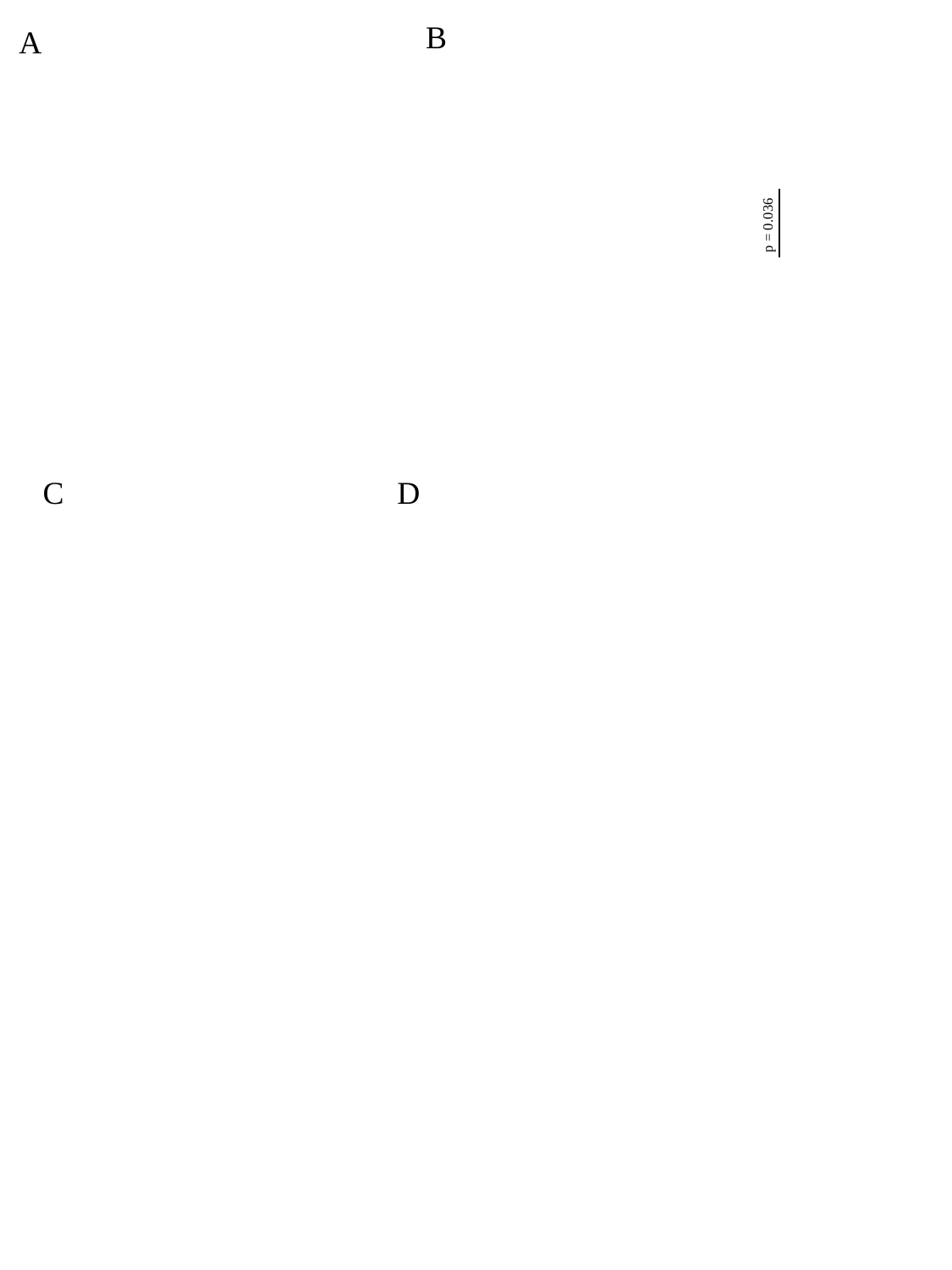

B
A
p = 0.036
C
D
